# Is large-scale rapid CoV-2 testing a substitute for lockdowns?

**DOI:** 10.1101/2021.04.26.21256094

**Authors:** Marc Diederichs, René Glawion, Peter G. Kremsner, Timo Mitze, Gernot J. Müller, Dominik Papies, Felix Schulz, Klaus Wälde

## Abstract

**BACKGROUND:** Various forms of contact restrictions have been adopted in response to the Covid-19 pandemic. Around February 2021, rapid testing appeared as a new policy instrument. Some claim it may serve as a substitute for contact restrictions.

**OBJECTIVES/METHODS:** We evaluate the effects of a unique policy experiment: In March and April 2021, the city of Tübingen set up a testing scheme while relaxing contact restrictions. We compare case rates in Tübingen county to an appropriately identified control unit.

**CONCLUSIONS:** The experiment led to an increase in the reported case rate. This increase is robust across alternative statistical specifications. An epidemiological model that corrects for ‘more cases due to more testing’ and ‘reduced testing and reporting during the Easter holiday’ also confirms the finding.

---

Can large-scale CoV-2 testing strategies substitute for restrictive public health measures? In theory, the idea is straightforward. If, first, every socially active person is subjected to a rapid CoV-2 test on a regular basis and, second, quarantined if tested positive, there is (almost) zero infection risk from social interactions. One would achieve the same outcome as under a complete lockdown—albeit at much lower costs: social interactions could be maintained.

In practice, there are several complications. Any testing procedure generates false negatives, that is, some infections will necessarily go undetected (*1*). Moreover, the timing of testing is critical: when testing takes place too early, infected persons go undetected, when it takes place too late, the transmission of the disease may have already taken place. Some therefore suggest that rapid tests do more harm than good (*2*). Lastly, testing and quarantining may be not sufficiently comprehensive, for instance, because of a lack of compliance.

Lockdowns, on the other hand, are also unlikely to prevent new infections altogether. First and foremost, they cannot be complete because some social interactions are essential. Second, their effectiveness also suffers from lack of compliance (*3, 4*).

Given this debate, an empirical assessment seems warranted. This paper turns to a uniquely suited policy experiment set up in the German town of Tübingen. Between March 16 and April 24, 2021, it ran a large-scale rapid testing scheme while simultaneously relaxing lockdown measures (appendix A.1). Each negatively-tested person was permitted, inter alia, to shop, go to movie theaters or join other people in restaurants (outdoors). While several towns tried to obtain similar permits elsewhere in Germany (*5*), the case of Tübingen is unique as its experiment started while other German counties were still in lockdown. We rely on these counties as a reference group in our synthetic control method (*6* –*9*) to assess whether largescale rapid CoV-2 testing can be a substitute for lockdowns. The answer would be yes if opening under safety did not increase cases in Tübingen.

## 1 Findings

### 1.1 Empirical findings

We describe the pandemic state by the key metric for policy decisions in Germany: the seven-day SARS-CoV-2 case rate (appendix A.2.1). The left panel in figure 1 shows the development of the case rate between February and April 2021. The solid black line in the left panel represents the development in Tübingen county, the dashed red vertical lines indicate the start and end of the policy experiment. The line for Tübingen county shows that the case rate was below 50 before the start of the project and increased to almost 150 during the Easter weekend starting April 2. This increase coincided with opening under safety (OuS) and led to wide public claims that “Tübingen failed”.

**Figure 1:**
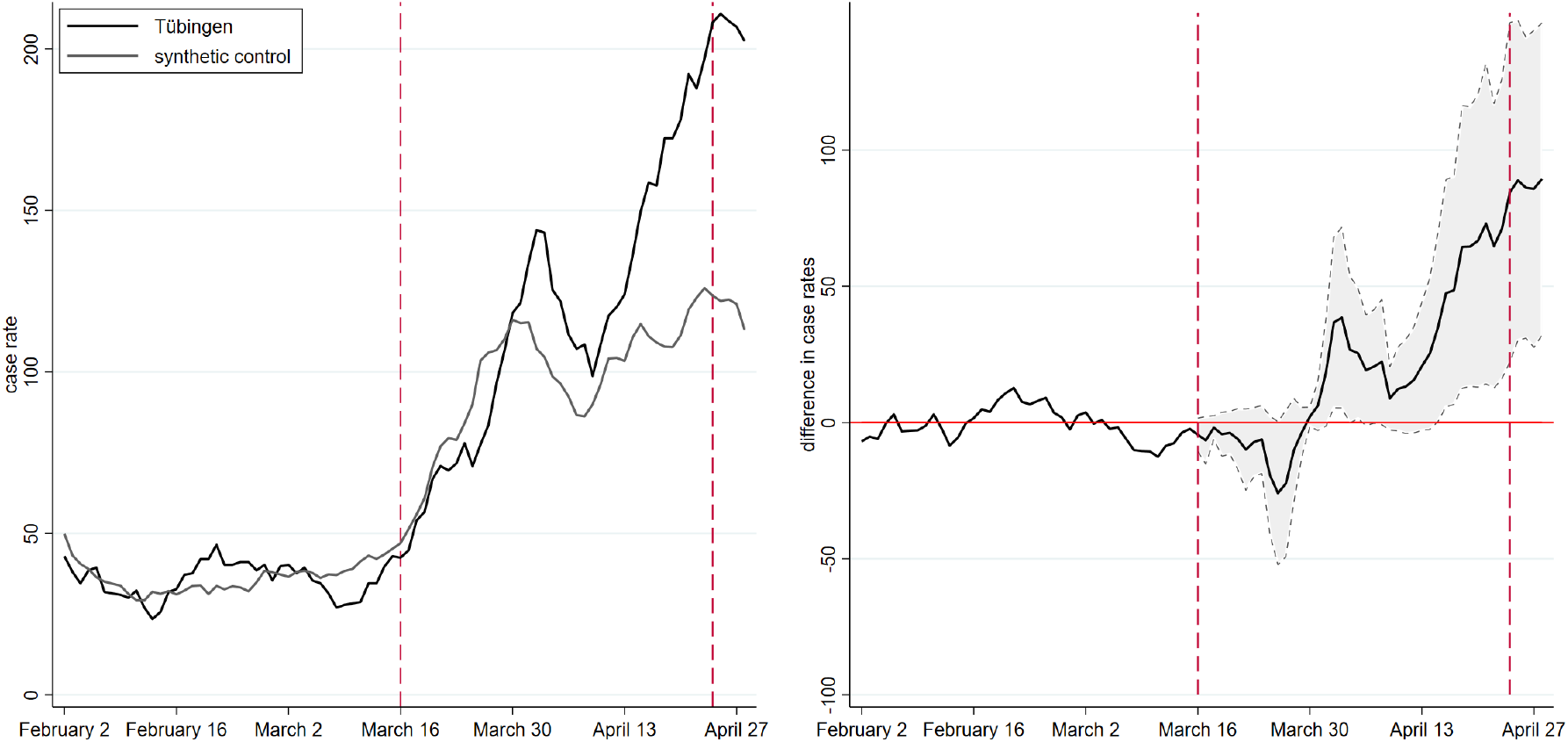
Seven-day case rates of Tübingen and control group

Statistics tells us, however, that we cannot assess the causal effect of a policy experiment by comparing the case rate before and after the start of the project. Other factors than OuS are likely to have affected pandemic dynamics in Tübingen over this period as well. We, therefore, need to compare the pandemic development in Tübingen county to a control group of similar counties: counties should display comparable pandemic dynamics before the start of OuS in Tübingen, should share certain fundamental socio-demographic and health care characteristics (e.g., population density, age structure, medical services, commuting patterns) and should be subject to very similar if not identical public health measures.

We identify such a set of control counties using the synthetic control method (see section 2). The resulting control counties and their weights constituting our synthetic control county are presented in table 1. The synthetic control county consists of four urban districts (‘Stadtkreis, SK’) and four rural districts (‘Landkreis, LK’). Two of the three units that receive the largest weights (Freiburg and Heidelberg) are cities in Baden-Württemberg with major universities that have similar population levels of up to 230K and comparable socio-demographic structures. Local health care systems are also similar. Table 5 in the appendix shows the details of the fit.

**Table 1:** Control counties and their weights for figure 1

| name | weight | name | weight |
| --- | --- | --- | --- |
| SK Freiburg i.Breisgau | 0.29 | LK Pfaffenhofen a.d.Ilm | 0.07 |
| LK Eichstätt | 0.24 | LK Bitburg-Prüm | 0.05 |
| SK Heidelberg | 0.19 | SK Münster | 0.04 |
| SK Oldenburg | 0.09 | LK Lüneburg | 0.03 |

Given this background, we can now again turn to figure 1. The solid black line, representing Tübingen county, and the grey line, representing the synthetic control, show very similar case rates prior to the beginning of the experiment, indicating a good fit in the pre-treatment period since February 2021. When we compare the development in reported case rates after the beginning of the experiment, we initially observe a parallel development between Tübingen county and its synthetic twin. At the beginning of April, cases in the control county start to decline, whereas the decline in Tübingen county only sets in a few days later. By April 10, the gap between Tübingen county and the control county is almost closed.

This development is also visible in the right panel of figure 1. A first peak in the difference between treatment and control occurs 2.5 weeks after the start around April 3, just around the Easter weekend. The right panel also shows that this difference, while visible, is hardly statistically different from zero at the 10% level. Nevertheless, a treatment effect is visible: OuS seems to increase the case rate – at least temporarily.

As of April 10, however, data not included in our earlier version (*10*), case rates in Tübingen more strongly increase relative to its synthetic twin. While some open questions related to OuS in Tübingen will be addressed in our discussion section, the most straightforward interpretation of this increase after April 10 is the continuation of a process OuS initiated on March 16: More contacts lead to more cases. The reduction in the gap between Tübingen and its synthetic twin is due to the Easter holidays and the slowdown of reporting of data from laboratories and doctors to local and national health authorities.

### 1.2 Testing and the Easter break

Any empirical finding calls for a theoretical interpretation. Empirically, we find that OuS increases case rates. Theoretically, at least two questions arise: Did case rates increase only because OuS implies more testing and when we test more, we find more? Second, can we believe our verbal interpretation that OuS and the Easter break imply such a non-monotonic behavior as visible in figure 1? We answer these questions in turn.

#### 1.2.1 Case rates and testing

Some argue that the number of reported infections increases when there is more testing. The argument is not convincing when a test is undertaken because a patient with Covid-19 symptoms visits a doctor. If doctors arrange for tests, the number of tests depends on the number of patients with Covid-19 symptoms. The number of reported infections therefore increases only when there are more patients with symptoms. Tests increase as a function of the state of the pandemic (*11*).

The argument is true when testing is the outcome of projects such as OuS. In this case, the number of tests does not depend on the state of the pandemic but on the number of participants that want to be tested. Similar arguments can be made with respect to testing travelers, testing sport professionals, or all other preventive testings. In this case, more infected individuals are found when there is more testing.

To understand the quantitative importance of this argument for OuS, we extend a standard SIR model in four respects (appendix A.5.2). We (a) allow for rapid testing leading to (b) discovery of asymptomatic (unreported) cases who, thereby, (c) turn into reported cases. We assume that (d) reported infectious individuals are in quarantine and infections can only occur when meeting a non-reported infectious individual (*12*). OuS in this framework consists of these four features plus an increase in the contact rate, i.e., as an example, the number of individuals one person meets per day.

When we want to separate the effect of more contacts from the effect of more testing, we first fit the extended SIR model to the data (appendix A.5.2). Second, we switch off the testing channel by assuming that no extra testing takes place: in this case, the model predicts that, all else equal, the increase in (reported) cases is less strong initially (appendix A.4.2). This finding supports the notion that more testing leads to more (reported) cases. However, the effect is quantitatively small (with a maximum of 7%) and vanishes over time because the increase in case rates accelerates in the absence of testing (and quarantining). Hence, the fact that more testing leads to more cases is unlikely to be the reason for the strong increase in the case rate in Tübingen (figure 1) in the context of OuS.

#### 1.2.2 OuS and Easter break

Our complete explanation of figure 1 builds on a combination of a (i) permanent OuS effect and a (ii) temporary Easter break effect. OuS has a permanent effect on the contact rate, the Easter break temporarily reduces reporting and testing. We capture these two effects by an additional feature of our SIR model (appendix A.5.2) that lets the flow from exposed to reported infectious individuals fall over Easter. A certain share of infectious individuals who display symptoms do not go to an emergency center and therefore do not get tested.

Employing this framework shows that one can easily explain the rise in case rates in Tübingen by an increase in contact rates (appendix A.4.3). The temporary drop over the Easter break can be understood by a reduction in transmission and testing. This finding holds when we estimate one unique increase of contacts and when we estimate two separate contact rates, one before and one as of Easter (table 6). We concede that the Easter effect in the SIR model is not as pronounced as in the data. What we can clearly see, however, is that the permanent effect of OuS can easily explain the entire increase in case rates. Our theoretical interpretation therefore confirms our empirical findings. OuS did increase case rates in Tübingen relative to its control group.

## 2 Method

### Implementing the synthetic control method

We estimate the causal effect of OuS (the ‘treatment’) on infection dynamics in Tübingen (the ‘treated unit’) by relying on the synthetic control method (SCM). It was proposed for the causal assessment of policy interventions based on aggregate outcome measures (*6, 7*). At the heart of this method lies an estimator which identifies, in our application, counties in Germany to which Tübingen county can be compared (the ‘synthetic control county’). This comparison is based on information observable prior to treatment and summarized by a set of predictor variables (the ‘predictor set’). SCM requires an a-priori list of counties (the ‘donor pool’) from which to construct the control unit. See appendix A.5.1 and table 5 for more background.

Results depend on how we measure the pandemic (the ‘outcome variable’). Our preferred outcome variable is the 7-day case rate. We also employ cumulative cases and, briefly, the positive rate, as alternatives. Robustness checks for the predictor set, the donor pool and outcome variables are undertaken and will be discussed shortly.

The present study puts special emphasis on two novel predictor variables. First, we allow for spatial controls. They are due to the low case rate of Tübingen compared to other counties in Germany before the start of OuS on March 16, visible in figure 4. If visitors enter Tübingen from counties with higher case rates, Tübingen will likely experience higher case rates itself. To this end, we looked for control counties that were also surrounded by counties with case rates similar to the neighbors of Tübingen. If Tübingen is subject to a ‘catching up’ process, we wanted to make sure that Tübingen is compared with regions that are also subject to ‘catching up’.

Second, it appears very important that counties are as similar as possible to Tübingen county in terms of Covid-19 policies. We achieved this goal in two ways. On the one hand, we constructed an index (see appendix A.2.4) that measures the stringency of Covid-19 policies. As an alternative, we compare Tübingen to counties from its state Baden-Württemberg only. Counties all coming from the same state as Tübingen are very homogeneous with respect to their Covid-19 rules.

### Estimating an extended SIR model

The differential effects of ‘opening’ and of ‘safety’ (i.e. testing) in OuS can be understood and quantified by estimating an extended SIR model (*13* –*15*). Our central extension of a standard SIR model (appendix A.5.2) consists in modeling rapid testing and the Easter break.

Testing implies a flow from asymptomatic, non-reported individuals to reported individuals. Assuming that all reported individuals enter quarantine, the share of infectious individuals in society (one can meet) falls due to testing. The Easter break implies that (i) test results are not reported to health authorities and that (ii) not all individuals with symptoms visit doctors.

We quantify the parameters of both extensions by matching cumulative cases, case rates and observed positive tests. We do so by minimizing the squared difference between data and model predictions (*14*). We infer the effects of testing on the reported number of infections by computing a hypothetical time series for cases under the assumption that no testing was undertaken in Tübingen. The effect of the Easter break is quantified by estimating the share of individuals that do not visit emergency units despite having symptoms.

## 3 Discussion

Findings from a comparison of a county with a synthetic county depend on (a) the measure used (outcome variable), (b) the criteria employed to find comparable counties (predictor set) and (c) the group of counties from which to choose comparable counties (donor pool). Varying our choices confirms the basic finding.

### 3.1 Predictor sets and donor pool

The outcome of our main robustness checks for Tübingen county are visible in figure 2. In addition to our predictor set ‘baseline’, employed for estimating results displayed in figure 1, we structure our discussion around two additional predictor sets, a predictor set 1 and a predictor set 2. Our baseline predictor set, visible in table 5, was discussed in the method section. Predictor sets 1 and 2 (visible in tables 14 and 16, respectively) shorten the pre-treatment period, starting on February 1 in the baseline predictor set, to a start date of March 1. This investigates into the importance of pandemic pre-treatment variables for the pre-treatment fit and for the overall result. Predictor set 1 employs daily case rates while predictor set 2 employs weekly case rates. Predictor set 2 therefore further reduces the importance of pre-treatment pandemic measures relative to non-pandemic variables. To understand the importance of spatial controls and of the stringency index, we employ a predictor set defined as the baseline predictor set without the stringency index and another one defined as baseline without spatial controls.

**Figure 2:**
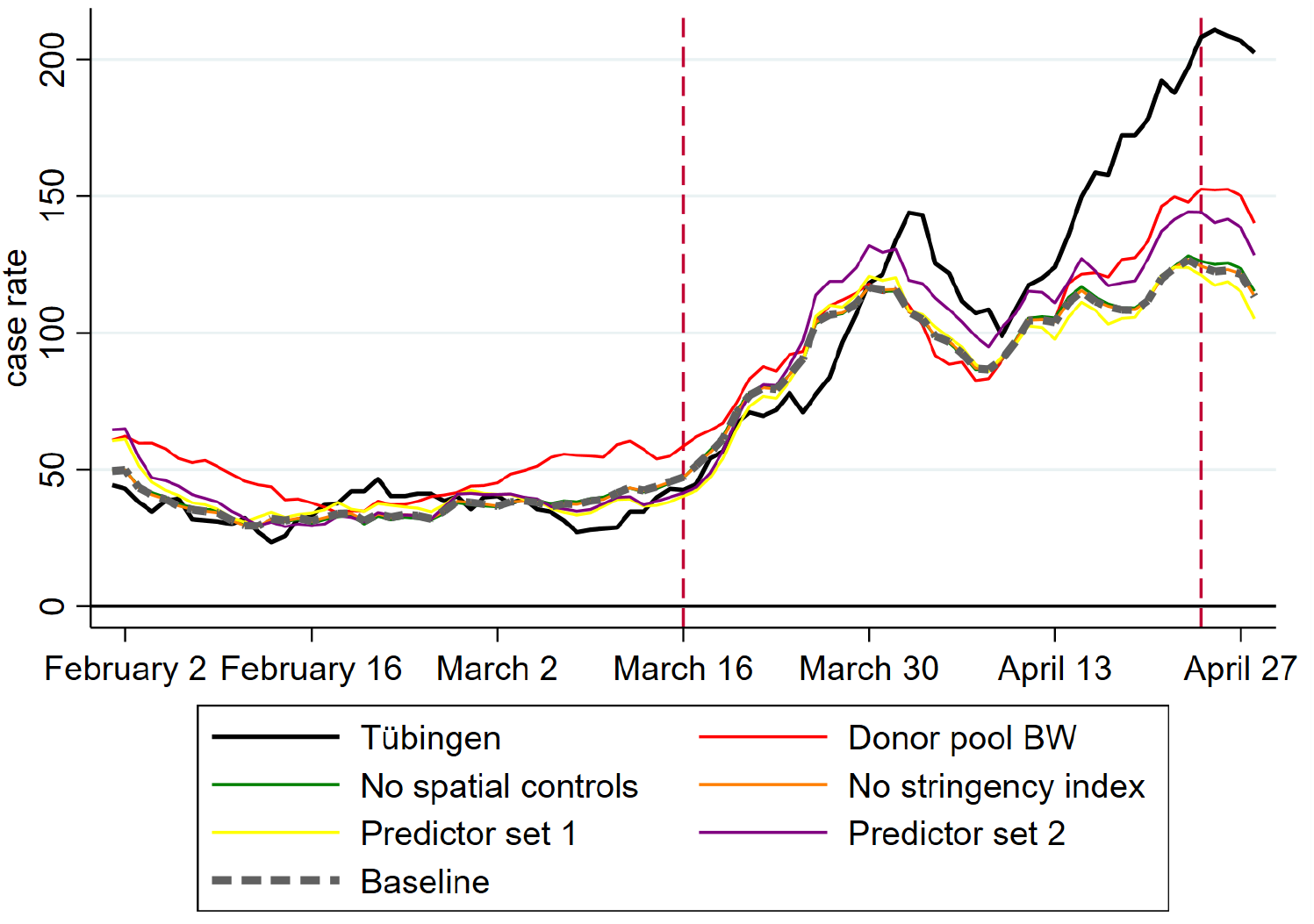
Seven-day case rates for alternative predictor sets and donor pools

As figure 2 shows, all of our robustness analyses confirm the baseline scenario. This is most impressively visible by the hardly visible (grey dashed) baseline graph in this figure: the predictor set without the stringency index leads to basically the same result. When we take out spatial controls, the prediction gets slightly better. This means that the ‘catching up’ argument laid out in the method section does not have a large quantitative importance. The shorter pre-treatment fitting period of predictor set 1 leads to a somewhat worse prediction than baseline. Again, quantitatively, this is of no importance.

Going from daily to weekly pre-treatment frequency with predictor set 2 worsens the pre-treatment fit but improves the post-treatment prediction. The same is true when we restrict the donor pool to Baden-Württemberg only. If we therefore put more emphasis on homogeneity in Covid-19 policy, we could tell a somewhat more optimistic story. Given the worse pre-treatment fit (RMSPE of 19.57 in table 10 instead of 9.9 in table 5), however, we do not put too much emphasis on this finding. We conclude that our baseline result is confirmed by these robustness tests that vary the predictor set and the donor pool.

#### 3.2 The role of the pandemic measure

The seven-day case rate as employed in figure 1 is the measure of the pandemic state that receives most of the attention around the world. It is not clear, however, whether this is the best measure for a pandemic. It is also not clear whether this is the best measure to compare the evolution of the pandemic across regions. A moving average over a period of seven days is much more short-run in nature than, for example, the sum of all new infections since some starting point.

We therefore employ the total number of reported infections since January 2021 per 100,000 inhabitants as dependent variable. Appendix A.7 shows that the synthetic twin of Tübingen consists of different counties than in our benchmark analysis. The fit dominates the baseline fit as cumulative infections over a longer period than only seven days are less volatile. Finding similar counties is therefore easier for the SCM. What is most important, however, is the evaluation of OuS: We confirm the findings from above. Interestingly, Tübingen and its control also move more or less in parallel up to around April 1. Only then, the difference becomes much larger, just as in the case of the case rates.

An alternative popular measure of the severity of a pandemic is the positive rate, i.e. the share of positive tests in the total number of tests being undertaken in a certain population. This was the official measure of the scientific team behind OuS in Tübingen (of which one of us was the head) and of policy-makers. By this measure, there was no increase in the severity of the pandemic in Tübingen. Local outbreaks in communities and residential homes might have increased the case rate in the county but were not visible in the positive rate.

#### 3.3 Is Tübingen city different?

Many commentators on OuS in Tübingen have argued that conclusions drawn from Tübingen county could be misleading. One should rather study Tübingen city. Probably findings on OuS would be less negative in this case, the claim goes.

The details of the timing of OuS in Tübingen are in table 2. All testing and opening measures took place in the city of Tübingen. As of April 6, only shopping in non-essential stores for local residents continued. Those parts of the experiments that attracted most visitors had ended. Maybe the increase in case rates in Tübingen county is due to increases in communities other than Tübingen city (see figure 17 for a map).

**Table 2:** Time of policy experiment

| Date | Change in permitted activities |
| --- | --- |
| March 16, 2021 | Opening of nonessential shops, outdoor dining, theaters, cinemas, etc. in Tübingen city center; official negative rapid test (“day ticket”) required |
| March 27, 2021 | Number of day tickets for visitors from outside Tübingen county limited to 3,000 per day |
| April 1, 2021 | Day tickets no longer available to visitors from outside Tübingen county |
| April 6, 2021 | Outdoor dining no longer possible |
| April 24, 2021 | Experiments ends |

To study this hypothesis, we first look at the evolution of case rates in Tübingen city, also shown in figure 4 in addition to Tübingen county. We observe that Tübingen city had indeed an (even) lower level of case rates than Tübingen county. We also see that Tübingen city followed the same overall trend as Tübingen county. What is more, figure 5 shows that *increase* in case rates in Tübingen city is larger than the increase in Tübingen county. This raises some doubts about the hypothesis.

To be on the safe side, we treat Tübingen city as an independent county and apply the SCM again. We adjust the predictor set (as different data are available at the community level than at the county level), the donor pool and the pandemic measure (see appendix A.9). We also adjust the pandemic measure as Tübingen city had one of the lowest, if not the lowest, case rates in all of Germany before the treatment date. This makes it impossible to construct an appropriate synthetic twin employing the case rate as pandemic measure. The new pandemic measure we use for Tübingen city is therefore the growth factor for case rates.

Before proceeding, we confirm that the use of a new predictor set and a new pandemic measure do not strongly affect our findings: We compare our baseline findings with findings based on the new predictor set and the new pandemic measure. Applying SCM then finally to Tübingen city shows that Tübingen city experienced a stronger increase in (growth of) case rates than its control regions.

Returning to the hypothesis, it is true that the increase in case rates in other communities belonging to Tübingen county contributed to the rise of cases in Tübingen county. There is just as much truth to the finding, however, that Tübingen city experienced a rise in case rates as well and that this rise is larger than the increase in comparable control counties.

## Data Availability

all data is public

## Acknowledgments

We would like to thank Peter Martus for guidance to testing data, Boris Palmer for support with data on Tübingen city and Oliver Piehl for general data support and background on official reporting of CoV-2 infections. For this research project, authors are solely funded by their universities, contributed equally and declare no competing interests.

## A Supplementary appendix

### A.1 The experiment

In order to appreciate the experiment under study, consider the developments in Germany prior to this experiment. German policy measures in response to the Corona pandemic are set at the state level. While policies differed somewhat across the 16 German states, all states agreed to a range of measures in response to the second wave in December 2020. In particular, non-essential shops, restaurants, and schools were closed. These measures were partly reversed in early March 2021 against the backdrop of rising infections numbers, presumably because the second wave of infections had died off by late February.

Tübingen is located in the state of Baden-Württemberg (BW, for short). Here, non-essential shops were opened on 8 March 2021, provided that the case rate in the county was below 50. Otherwise, a ‘click & meet’ scheme was put in place (i.e., shopping was permitted for customers with appointment). Teaching at primary schools resumed on March 15. These measures were announced on March 5 by the state government and hence implemented on short notice.

While regulations in Tübingen were mostly the same until then, the state government announced on March 15 that starting the next day (see table 2), the town of Tübingen would embark on a special experiment, centered around a large-scale rapid testing scheme, officially labeled ‘Opening under Safety’ (‘Ö ffnen mit Sicherheit’). The town set up 9 testing posts where everybody could be tested with a rapid antigen test free of charge. The capacity for daily testing was 9000 and there were more than 30K tests per week (*1*). 15 minutes after the test, the result would be released and in case it was negative, the subject was provided with a ‘day ticket’ entitling the holder to shop in non-essential stores, attend bars and restaurants (outdoors), cinemas and theaters (the OuS activities). In case the test was positive, people were asked to take a PCR test. A positive PCR test result is automatically reported to the public health office (‘Gesundheitsamt’). The PCR tests form the basis for the official statistics on which our analysis is based. OuS ended on Saturday, 24 April, due to a change in the ‘Bundesinfektionsschutzgesetz’ adopted by the German federal parliament.

Below the level of the 16 states, Germany is subdivided into a total of 401 counties (“Land-kreise” and “kreisfreie Städte”). Tübingen city (pop: 91K) is part of Tübingen county (pop: 229K). In total, there are 44 counties in BW. The experiment under study took place in Tübingen city only. Still, everyone living in Tübingen county was allowed to participate. Hence, spillovers from the city to other areas of the county may have potentially been significant. Our main analysis therefore focuses on a comparison of Tübingen county to other counties. Also, detailed data is available only at the county level.

To measure the causal impact of OuS, it is important to note that Tübingen is not exceptional in terms of fundamentals. However, it performed relatively well compared to its BW peers regarding CoV-2 case numbers (see appendix A.6.2 for more background). At some point, Tübingen county was indeed enjoying the lowest case rate in all of BW. Still, there have been many counties which did similarly well during the period. The experiment taking place in Tübingen rather than elsewhere is most likely due to local idiosyncrasies and politics that are orthogonal to infection dynamics. The experiment, while approved by the state government, was devised jointly by the town’s major and the Corona commissioner of Tübingen county. Both have gained prominence in national media as a result of vocal and eloquent interventions regarding the handling of the pandemic and, more importantly, because of their personalities. It seems that these personalities, rather than any special developments in Tübingen, have been causal for setting up the Tübingen experiment. It thus comes close to a randomized control trial.

### A.2 Data

#### A.2.1 General information

##### Cases, cumulative cases and case rates

To avoid confusion, it is useful to remind us of definitions of (cumulative) cases and case rates. The number of SARS-CoV-2 cases reported on a day *t* is given by the number of new infections on day *t* — 1. Cumulative cases over the previous *d* days on day *t* is given by the sum of cases from *t* − 1 − *d* to *t* − 1. The seven day case rate on day *t* is given by cumulative cases over the previous 7 days relative to *t*, divided by population size and multiplied by 100K.

##### County-level data

Data on reported SARS-CoV-2 infections at the county level are taken from the Robert Koch Institute (*2*). Infections are identified by PCR tests. For our empirical analysis, we use aggregate case numbers for each county and day based on the reporting date by local health authorities. Time-varying predictors are the average daily temperature and daily mobility changes for each county during the pre-treatment period until March 16, 2021. Mobility changes (in percent) based on individual mobile phone data are computed as the difference in mobility patterns between a specific date and the average value for the corresponding weekday from the same month in 2019 (pre-COVID benchmark period). To give a specific example: The mobility change for Wednesday, March 10, 2021, is calculated as the difference in the number of regional trips for this date and the average number of trips on Wednesdays in March 2019. We use data on daily temperatures from Deutscher Wetterdienst (*3*), and updated data on mobility changes per county and day are obtained from (*4*).

We further include time-constant cross-sectional predictors characterizing regional demographic structures and the regional health care system as in (*5*) based on data from the INKAR online database of the Federal Institute for Research on Building, Urban Affairs and Spatial Development (*6*). We use the latest year available in the database, which is 2017, and rely on the following cross-sectional predictor variables: population density (population/km^2^), the share of females in the population (in %), the average age of female and male population (in years), old- and young-age dependency ratios (in %), the number of medical doctors per 10,000 of population and pharmacies per 100,000 of population, the regional settlement structure (categorical dummy), and the share of highly educated population (in %).

##### Community-level data

To supplement our county-level-analyses with an analysis at the city level, we obtained daily case rates for the city of Tübingen for the treatment period directly from the city’s Corona Commissioner. For the pre-treatment period, we rely on weekly case rates that were provided to us by the local health authorities (https://www.kreis-tuebingen.de/17094149.html).

##### SCM data and repository

Data used for our SCM-based analysis fall into four groups: Data for our main county analysis, for the Tübingen city analysis, for the stringency index (section A.2.4) and some rapid test data (displayed in figure 3).

**Figure 3:**
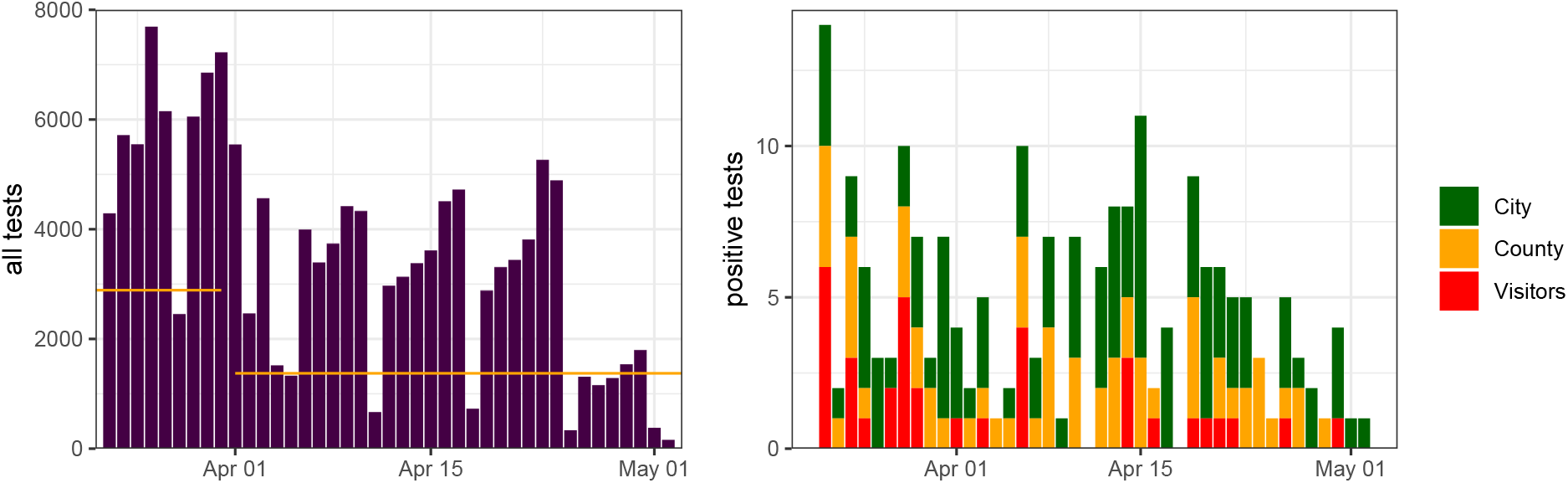
Total number of tests (left) and positive tests by origin of tested individual (right)

All of these data plus the corresponding Stata, R and matlab scripts are available in a public repository at https://figshare.com/s/87f712ecb2f9eaf044b1.

#### A.2.2 Descriptive statistics

Table 3 shows descriptive statistics for variables we employ for our main analysis. The variables are measured at the county level and the underlying population is Germany without direct neighboring counties of Tübingen (which are Böblingen, Esslingen, Reutlingen, Zollernalb, Freudenstadt and Calw). The latter are excluded from all analyses. Panel A contains all variables related to measuring the development of the pandemic. Panel B displays information on the time-varying predictors, mobility and average air temperature, and panel C shows all predictors related to the county’s demographic structure and their health care coverage.

**Table 3:** Descriptive statistics

|  | Mean | S.D. | Min. | Max. |
| --- | --- | --- | --- | --- |
| A: Data on reported CoV-2 cases |  |  |  |  |
| Seven-day CoV-2 case notification rate per 100,000 | 116.53 | 72.60 | 3.74 | 663.76 |
| Cumulative infections per 100,000 inhabitants since January 1st | 6290.96 | 8803.45 | 387 | 164461 |
| Cumulative cases over previous 7 days | 232.10 | 309.93 | 2 | 7340 |
| Cumulative cases over previous 14 days | 448.63 | 589.31 | 7 | 13428 |
| Neighbourhood (50km) seven-day case rate per 100k | 116.01 | 58.55 | 0 | 552.48 |
| B: Time-varying predictors |  |  |  |  |
| Average mobility | -.10 | .13 | -.69 | .73 |
| Average temperature | 3.70 | 4.81 | -17.5 | 19 |
| Stringency index | 2.88 | .18 | 2.38 | 3.16 |
| C: Regional demographic structure and local health care system |  |  |  |  |
| Population density (inhabitants/km<sup>2</sup>) | 535.44 | 705.39 | 36.13 | 4686.17 |
| Share of females in population (in %) | 50.60 | .64 | 48.39 | 52.74 |
| Average age of female population (in years) | 45.88 | 2.12 | 40.70 | 52.12 |
| Average age of male population (in years) | 43.18 | 1.84 | 38.80 | 48.20 |
| Old-age dependency ratio (persons aged 65 years and above per 100 of population aged 15-64 years) | 34.39 | 5.49 | 22.40 | 53.98 |
| Young-age dependency ratio (persons aged 14 years and under per 100 of population aged 15-64 years) | 20.53 | 1.44 | 15.08 | 24.68 |
| Medical doctors per 10,000 of population | 14.62 | 4.42 | 7.33 | 30.48 |
| Pharmacies per 100,000 population | 27.04 | 4.91 | 18.15 | 51.68 |
| Categorical variable<sup>§</sup> for population density of NUTS3 region | 2.60 | 1.05 | 1 | 4 |
| Share of highly educated* persons in regional population (in %) | 13.05 | 6.21 | 5.59 | 42.93 |

Notes: \* = International Standard Classification of Education (ISCED) Level 6 and above; <sup>§</sup> = included categories are 1) larger cities (kreisfreie Großstädte), 2) urban districts (städtische Kreise), 3) rural districts (ländliche Kreise mit Verdichtungsansätzen), 4) sparsely populated rural districts (dünn besiedelte ländliche Kreise).

A day-by-day overview of the number of tests and positive cases is provided in figure 3. On April 1, participation in the experiment was restricted to inhabitants of Tübingen county. Accordingly, we find a structural break in the data on this date with around twice as many tests being administered before (on average 2888 per day) than after the restriction (1373 daily tests). We are able to differentiate the number of positive tests into participants from Tübingen city, Tübingen county and elsewhere. We point out that the number of positive tests taken by visitors from the county after April 1 can be explained by commuting staff that was also frequently tested.

#### A.2.3 Tübingen and its donor pool

The SCM selects control counties from a donor pool. When we compare Tübingen to all regions in the donor pool, we get a first idea about the trend of Tübingen relative to other counties and about the highest and the lowest possible treatment effect. Figure 4 plots case rates of Tübingen county and Tübingen city within case rates of all German counties in the left panel. The right panel shows the same two time series within case rates of all counties from Baden-Württemberg.

**Figure 4:**
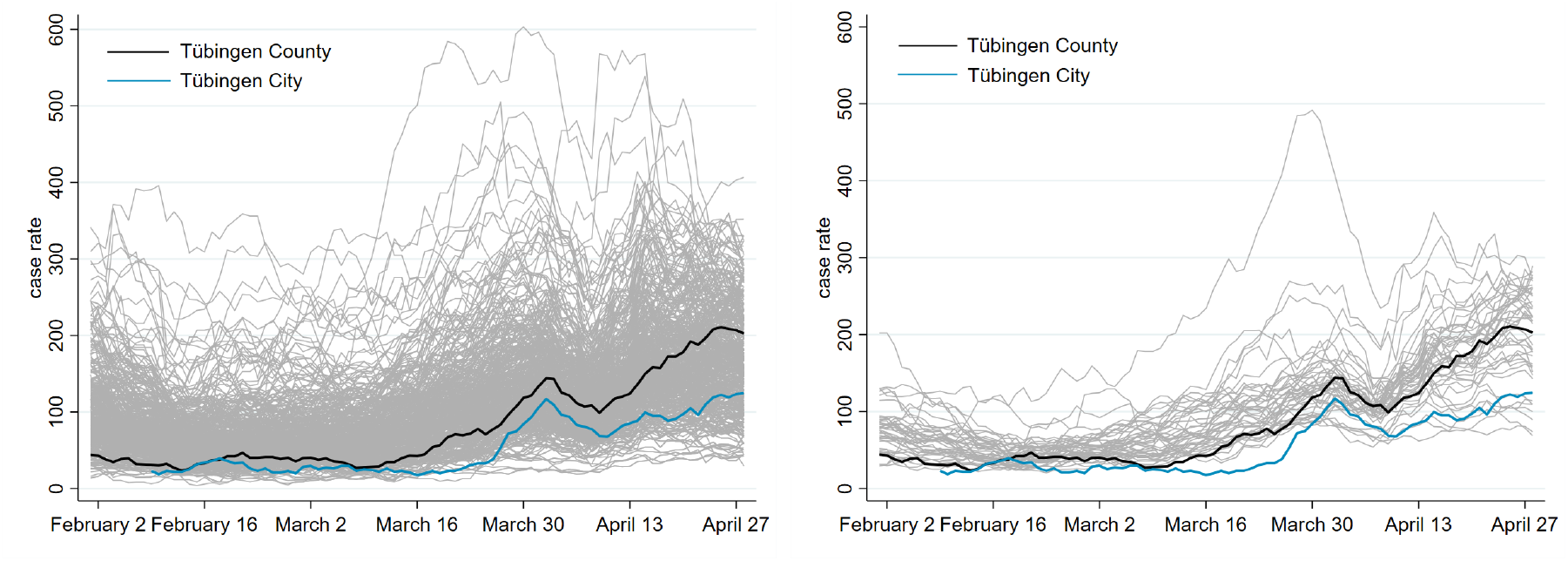
Seven-day case rates in Germany (left) and Baden-Württemberg (right)

Judging the effect of OuS from these figures is difficult for many reasons. One reason is that treated and control regions start from different levels. Figure 5 therefore normalizes the case rate on the treatment date (16 March) in all counties to 1. One can then directly read from the resulting figure whether the growth process in the treated regions was stronger over the treatment period than in control regions.

**Figure 5:**
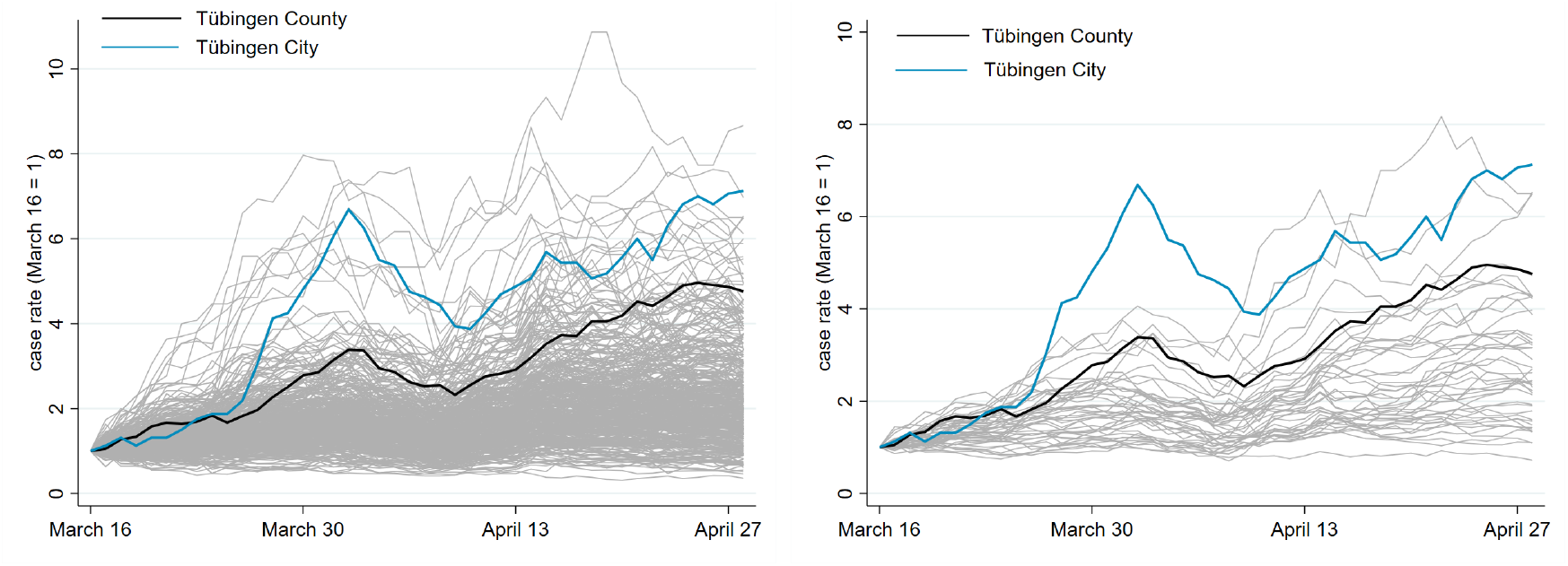
Normalized seven-day case rates in Germany (left) and Baden-Württemberg (right)

While these figures, of course, also do not allow to draw any causal conclusions about OuS, the relative increase of Tübingen city to Tübingen county came as a surprise. While Tübingen city always has a lower incidence (level), its incidence growth is much more pronounced, both relative to Tübingen county and relative to all counties in Baden-Württemberg.

#### A.2.4 A stringency index for German states and counties

Understanding the effects of almost anything related to the pandemic requires a detailed understanding of the institutional environment. Any region is subject to a long list of regulations that govern contacts in the private, in the public and on the workspace. In our analysis, we account for the largely decentralized policy framework enacted by German states and counties. Ideally, our synthetic control region consists of counties with a regime similar to pre-OuS conditions in Tübingen. To this end, we construct an index of the stringency of health regulations, similar to previous efforts based on an ordinal classification of measurements (*7, 8*). Building on the Infas database (*9*) and prior experience in this field of research (*10*), we are able to observe differences across counties.

The available data allows us to distinguish between *k* = 1…*K* domains like e.g. kindergartens, shops and restaurants. Up to 16 distinct policies are documented for each domain. In line with the other indices, we group the policies by strictness into five levels from 0 to 4. Hence, for each region *i*, day *t* and domain *k*, there is a vector *L*_*i,t,k*_ with 16 values. The highest of these 16 values is given by max *L*_*i,t,k*_. The index *I*_*i,t*_ ∈ [0, 4] is then calculated as daily averages of the maximum value of the *K* = 23 domains,

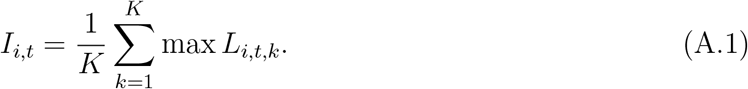

A first impression of the stringency of policies over time is given in figure 6. We plot the index *I* on the vertical and time on the horizontal axis. Each of the German states is represented using an individual color, Tübingen county is highlighted as the black line. As the figure reveals, policies are very homogeneous across counties. While there are 401 counties in Germany, there are rarely more than 20 different values visible at any point in time.

**Figure 6:**
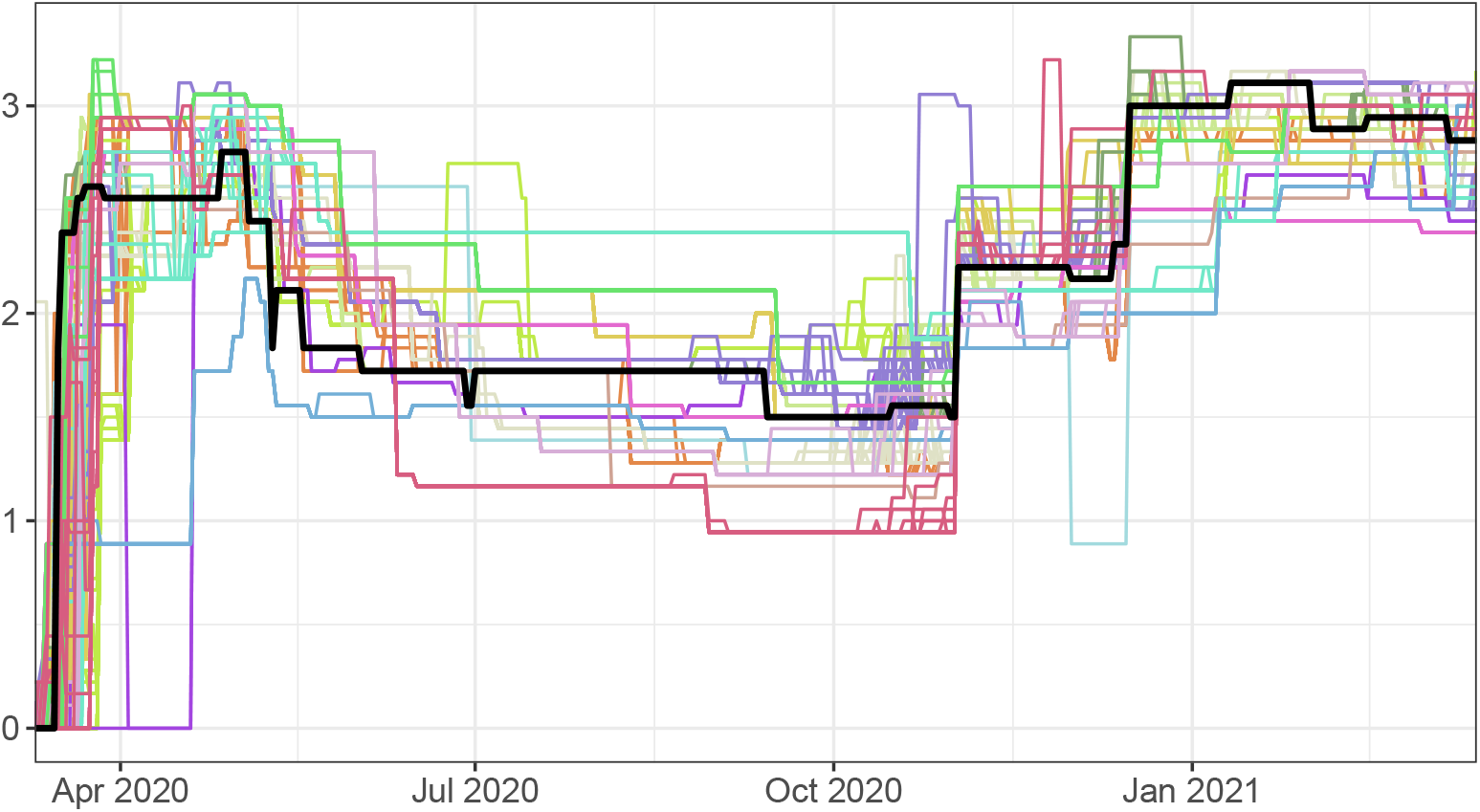
Stringency index across all 401 German counties.

For our SCM, we include the index in the pre-treatment period in the predictor set. This provides us with a control group that enforced measurements of similar scope and severity prior to the experiment as Tübingen did. We cannot use the index, however, to detect other OuS projects across Germany. This is due to the focus of the Infas database on county legislature. OuS projects are in most cases restricted to single communities and in many cases not mentioned in the county legislature. This makes it impossible to point out relevant regions based on the index. We therefore proceed with an exclusion of counties based on our compilation of OuS projects across Germany in table 4. The source for this search is general public information. We make sure that none of these counties appears in any of our synthetic control counties.

**Table 4:** Opening under Safety projects in Germany as of April 23, 2021

| Community | County | Start | End | Source |
| --- | --- | --- | --- | --- |
| Tübingen (City) | Tübingen | March 16 | April 23* | tuebingen.de |
| Weimar (City) | Weimar (City) | March 29 | March 31 | tmasgff.de |
| Augustusburg | Mittelsachsen | April 1 | April 23* | augustusburg.de |
| Nordhausen | Nordhausen | April 6 | April 16 | mdr.de |
| Alsfeld | Vogelsbergkreis | April 8 | April 15 | alsfeld.de |
| several | Harz | April 9 | April 23* | kreis-hz.de |
| Baunatal | Kassel | April 12 | April 23* | baunatal.de |
| several | Schleswig-Flensburg | April 19 | ongoing | ostseefjordschlei.de |
| several | Rendsburg-Eckernförde | April 19 | ongoing | ostseefjordschlei.de |
| all counties in the state of Saarland |  | April 6 | April 23* | saarland.de |

*Note:* All end dates marked with a \* have ended due to the enactment of unitary federal restrictions on April 24 (11)

### A.3 Literature

There have been calls for comprehensive and large-scale testing schemes early in the pandemic (*12*). In theory, it is clear that testing and quarantining can dramatically reduce the costs of an epidemic (*13*). A systematic empirical assessment, however, of the benefits of widespread rapid testing based on antigen tests is still missing (*14*). In the present paper, we contribute to such an assessment by studying a unique policy experiment in which widespread rapid antigen tests were coupled with opening of non-essential infrastructure. We estimate the causal effect of this intervention using the synthetic control method (*15* –*17*). This method, SCM for short, is the vehicle for our empirical identification strategy.

SCM has been frequently used in the social sciences to study the effect of policy interventions, broadly defined, on political, social, and economic outcomes (*17*). In these contexts, SCM has been shown to be a flexible and robust estimation tool. In addition, it has also been applied to COVID-related research, for instance, to study the effectiveness of lockdown measures by means of a counterfactual analysis for Sweden (*18, 19*) and to study the effect of shelter-in-place policies in California (*20*). (*10*) use SCM to study the effect of face masks on SAR-CoV-2 cases in Germany. The SCM approach was also used in the interim evaluation of the Liverpool mass-scale testing project (*21*). Similar to the Tübingen experiment, this pilot was centered around repeated testing of asymptomatic individuals. Those with a negative result were not allowed, however, to participate in otherwise restricted activities. Compared to the synthetic control region, they find that large scale testing does not significantly decrease case numbers and hospitalization. In a different context, SCM allowed quantifying the impact of the Brexit referendum on economic performance in the UK (*22*).

### A.4 Findings

#### A.4.1 Our baseline result

The synthetic twin county employed in figure 1 consists of control counties who are listed, jointly with their weights, in the main text in table 1. Table 5 below displays the criteria (predictors) which serve as basis for constructing the synthetic twin. Predictor values pertain to the pre-treatment period ending March 16, 2021.

**Table 5:** Balancing properties of predictor set ’baseline’ for figure 1

|  | Treated | Synthetic |
| --- | --- | --- |
| Seven-day case rate per 100k (Feb 1) | 44.30 | 49.48 |
| Seven-day case rate per 100k (Feb 8) | 31.45 | 34.44 |
| Seven-day case rate per 100k (Feb 15) | 31.89 | 32.19 |
| Seven-day case rate per 100k (Feb 22) | 40.31 | 32.70 |
| Seven-day case rate per 100k (Mar 1) | 39.87 | 37.29 |
| Seven-day case rate rate per 100k (Mar 8) | 27.02 | 37.12 |
| Seven-day case rate per 100k (Mar 15) | 42.97 | 45.26 |
| Cumulative cases over previous 7 days (Mar 9) | 63.00 | 72.70 |
| Cumulative cases over previous 14 days (Mar 15) | 158.00 | 150.28 |
| Average mobility (Mar 9 - Mar 15) | 0.00 | -0.16 |
| Average Temperature (Mar 9 - Mar 15) | 4.21 | 5.88 |
| Population density | 434.86 | 945.33 |
| Share of females in population | 51.26 | 51.06 |
| Average age of female population | 41.67 | 41.91 |
| Average age of male population | 40.03 | 39.98 |
| Old-age dependency ratio | 24.58 | 24.97 |
| Young-age dependency ratio | 20.20 | 19.73 |
| Medical doctors per population | 15.64 | 19.76 |
| Pharmacies per population | 23.48 | 27.42 |
| Categorical variable for population density of NUTS3 region | 2.00 | 1.86 |
| Share of highly educated persons in regional population | 26.47 | 26.09 |
| Stringency Index | 2.97 | 2.96 |
| Neighborhood (50km) seven-day case rate per 100k (Feb 1) | 81.64 | 80.99 |
| Neighborhood (50km) seven-day case rate per 100k (Feb 8) | 61.83 | 63.29 |
| Neighborhood (50km) seven-day case rate per 100k (Feb 15) | 51.14 | 49.45 |
| Neighborhood (50km) seven-day case rate per 100k (Feb 22) | 44.51 | 47.05 |
| Neighborhood (50km) seven-day case rate per 100k (Mar 1) | 58.36 | 49.55 |
| Neighborhood (50km) seven-day case rate per 100k (Mar 8) | 56.70 | 54.15 |
| Neighborhood (50km) seven-day case rate per 100k (Mar 15) | 70.73 | 72.95 |
| RMSPE (pre-treatment) | 9.90 |  |

*Note:* Dates in parentheses indicate when the respective variable was measured.

Predictors can be split into groups: lagged pandemic measures (the outcome variable) and structural regional characteristics, which are expected to influence the local infection dynamics over time. As the table shows, we place a strong emphasize on lagged values of the seven-day case rate as predictor in order to ensure that Tübingen and the selected control regions follow a common pre-trend in the last two weeks before the OuS experiment stated in Tübingen. We also include an average measure for the cumulative number of SARS-CoV-2 cases in the two weeks before treatment start.

It would have been nice to include data on (UK) mutant shares. Unfortunately, they are not available at the county level. We (implicitly) capture mutant effect by focusing on pre-treatment pandemic dynamics when selecting control regions.

With regard to structural regional characteristics, we use both time-varying and time-constant predictors. As such, we use average levels for daily temperature and intra-regional mobility changes in the week prior to the treatment. The link between seasonality and infection dynamics has recently been studied (*23*). Including mobility effectively controls for social interaction as a driver of local infection dynamics and also as a measure how closely people follow prevailing (lockdown) policy rules (*24*).

Additionally, we control for the share of females in population, average age of female population, average age of male population, old-age dependency ratio, young-age dependency ratio, medical doctors per population, pharmacies per population, categorial variable for population density of counties and share of highly educated persons in regional population as suggested in (*5*). The rationale behind the inclusion of these predictors is to match Tübingen as closely as possible to its synthetic control group in terms of socio-demographic factors and factors related to the local health care system. Previous research has shown that these factors are significantly related to differences in COVID-19 incidence and death rates at the sub-national level (*25*).

The fit of the weighted predictor variables of the control counties from table 1 with respect to corresponding variables in Tübingen can be seen from comparing column 2 and 3 in table 5. Overall, there is a good fit. The population density is roughly twice as high in the synthetic control group as in Tübingen. We do not believe, however, that this is crucial for our findings. If anything, our effects are underestimated as the speed at which infection spread should be higher in more densely populated counties. The overall good fit underlines the good pre-treatment fit between the seven-day case rate development in Tübingen and its synthetic control group, as already visualized in figure 1.

#### A.4.2 Separating the effect of opening and the effect of ’safety’ (testing) in OuS

Section A.5.2 presents an extended SIR model which allows us to disentangle the effect of testing from the effect of opening and to understand the effect of the Easter break. The following section shows the effect of testing while the subsequent section A.4.3 focuses on the effects of the Easter break.

##### Distinguishing testing from opening

We use the baseline calibration (figure 11) to compute the evolution of the case rate in case of no testing. We simply set the detection rate equal to zero, *λ*^test^ = 0, as of *t*=March 16 and solve the model again.

Figure 7 considers the period from the beginning of OuS (16 March) to its end (24 April). The left panel shows the case rate under OuS as of March 16 (black graph). The black graph captures the general pandemic dynamics, the effect of more contacts and the effect of more testing, i.e. the general pandemic plus OuS. The solid graph is the best fit of the model case rate to observed data (see top right panel in figure 11).

**Figure 7:**
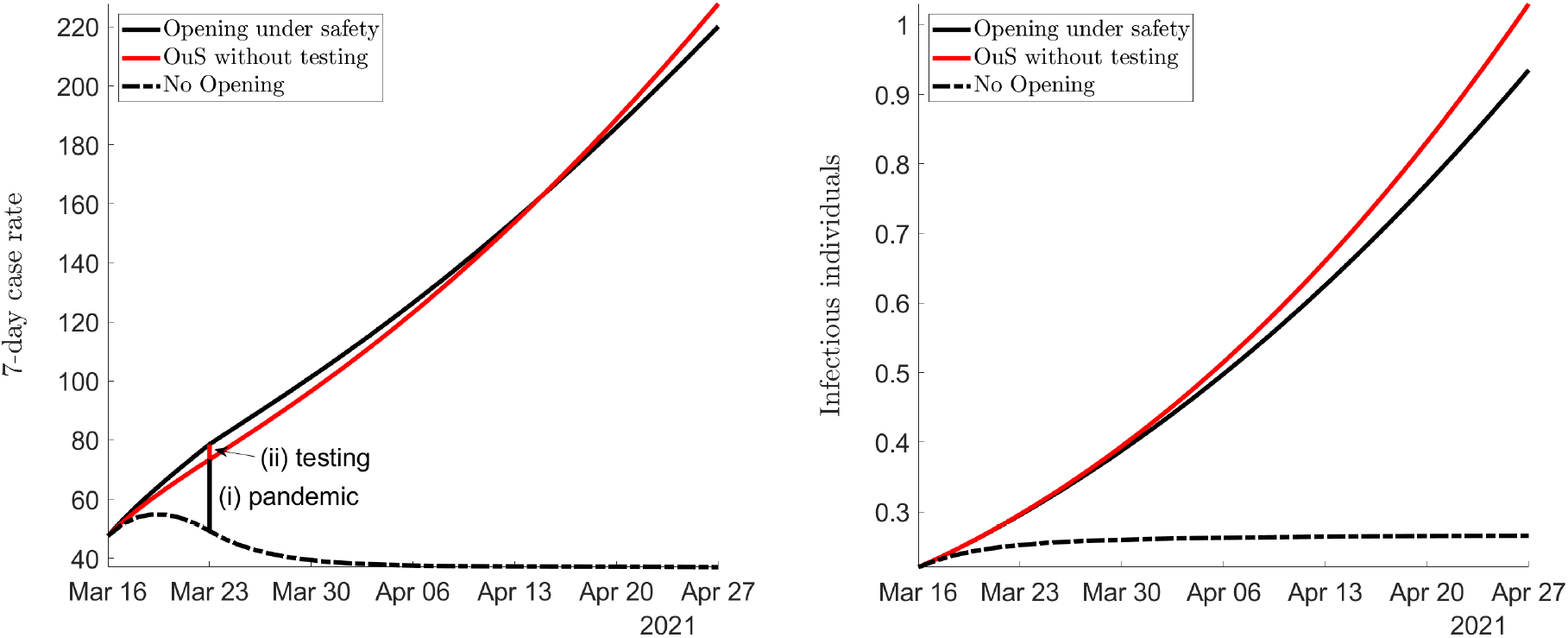
The effect of testing on case rates (left) and the true pandemic state (right)

The counterfactual scenario of no testing (red graph) yields case rates that would have been observed if only opening had taken place, i.e. if only (a) the contact rate had increased in Tübingen but without (b) the identification of asymptomatic infected individuals and without (c) individuals entering quarantine. Comparing this counterfactual scenario to baseline shows that case rates would have been lower initially but then higher as of April 15. The part due to testing (consisting of (b) and (c) and shown as (ii) in the figure) on case rates can be positive (which is (b): more testing leads to more reporting) or negative (which is (c): more testing puts more infectious individuals into quarantine). The example highlighted in the figure therefore qualifies the statement that more testing leads to more reported cases. The figure also highlights the difference (i) between the control region and Tübingen due to the pandemic effect of OuS.

Let us now turn to the true pandemic state, defined as (the share of) individuals that are infectious (reported or not). The right panel shows that the number of infectious individuals under rapid testing (black) falls as of the moment testing starts relative to the number of infectious individuals without rapid testing (red). Testing is obviously always good for the true pandemic state but not necessarily so for the measured pandemic state (i.e. reported case rates). Quantitatively, the increase of case rates due to testing in Tübingen is very small. When we divide the black by the red graph (left panel), the maximum increase in the case rate due to rapid testing is below 7% (see bottom right panel in figure 11).

This analysis shows that we should correct reported case rates by the factor found in the bottom right panel in figure 11. We would obtain a time series for revised case rates that reflects the effect of opening only, but not the effect due to more testing. This is point (i) above. The corrected cases (i) are then the appropriate time series for Tübingen that should be compared to its control unit. Looking at these graphs as of April 15, however, the case rate is actually lower in the case of OuS. As of April 15, we therefore need to *increase* reported numbers. On April 27, case rates under OuS without the testing effect would have amounted to 226 instead of 219 under OuS. Observing a maximum initial correction of 7% and the requirement to increase case rates as of April 15, it is clear that we would reach the same conclusion about OuS in Tübingen as in our baseline analysis in figure 1.

##### Increasing the testing pace

To illustrate the principle effect of testing even more clearly, consider figure 8. It replicates the findings just reported (OuS and OuS without testing) and asks what the effects would have been if testing had been higher by a factor of 5 (yellow curve) or even 10 (green curve).

**Figure 8:**
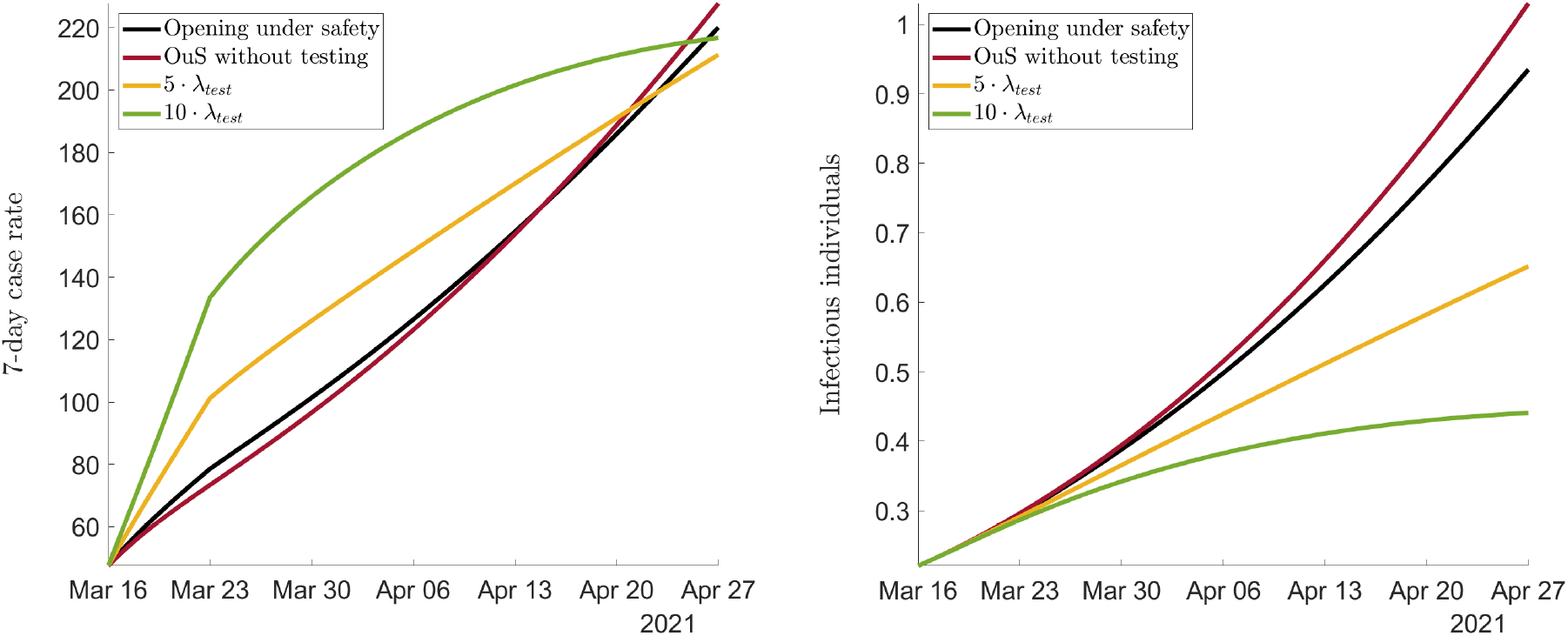
The effect of increasing the number of rapid tests

One can see the discrepancy between what is measured (the case rates in the left panel) and the true pandemic state (in the right panel) most clearly with the green graph. When we test ‘a lot’ (i.e. 10 times more than what actually took place in Tübingen), the reported case rate goes up a lot (left), yet, the true pandemic state goes down strongly (right). Again, the argument that more testing leads to higher cases is confirmed, but only initially. After around 6 weeks, the number of reported cases is lower even when testing ‘a lot’. As the right panel shows, the more is tested, the lower individual infections. This again stresses the discrepancy between the true state of the pandemic (right panel) and the measured state (left panel).

##### An ‘intuitive’ data correction

In earlier work (*26*), we corrected observed case rates by the number of positive tests in rapid test centers (adjusted for a factor taking false positive tests into account). To understand why this more intuitive approach of correcting for “more cases due to more testing” is theoretically not consistent, we go back to our true pandemic state, the number of infectious individuals, shown in the right panel of figure 7. When we take the theoretical tradition in epidemiology, as cast into various versions of the SIR model, seriously, we can ask whether subtracting positive tests from observed cases (analysing test *rates* and case *rates* would lead to the same insight) leads to a measure which is related to the true pandemic state. Given our SIR model (A.4) - (A.10), reported cases are given by *I*_*r*_(*t*). When we remove the number of positive tests *λ*^test^*I*_*n*_(*t*) from the inflow into *I*_*r*_ in (A.6), we do get some corrected number of cases. It would be described by the ODE 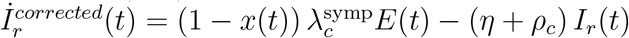.

This ‘intuitive’ correction does not lead to a measure of the true pandemic state, however, as any reference to asymptomatic cases *I*_*n*_(*t*) is missing. We therefore stick to our theoretically better founded correction displayed in figure 7.

#### A.4.3 OuS and Easter break

We understand our SCM findings such that OuS increases contacts and thereby case rates, even though additional testing leads to the detection of more cases, and the Easter break leads to a temporary dip in case rates. We would now like to understand whether our extended SIR model can support this interpretation. In addition to quantifying the effect of testing, we now also take the effect of the Easter break into account. To this end, we estimate the share of individuals that do not consult medical emergency services during public holidays.

All calibrated parameters are collected in table 6. The table shows the contact rate *a* and parameters *α* and *β* from our specification of the infection rate (A.11), the detection rate *λ*^test^ with which asymptomatic individuals are identified and the share *x* of symptomatic individuals that do not visit health centers during holidays. See the illustration of our SIR model in figure 10 and the full SIR model in (A.4) - (A.10).

**Table 6:** Calibration Results for the SEIR model

| Region | $a$ | $\alpha$ | $\beta$ | $\lambda^{\text{test}}$ | $x$ |
| --- | --- | --- | --- | --- | --- |
| OuS only | .0025 | .0027 | .63 | .026 | — |
| Control | .0016 | .1764 | 6.1e-07 | 0 | — |
| OuS & Easter throughout | .0027 | .0114 | .7118 | .0115 | .3335 |
| OuS & before Easter | .0029 | .0288 | .6986 | .0132 | — |
| OuS & as of Easter | .0021 | 4.1e-06 | .5602 | .0132 | .3944 |

##### Testing only

When we estimate the testing effect only and neglect the Easter Break, we get the first line in table 6 for Tübingen. The corresponding values for the control county is in the second line. These values were used above to plot figures 7, 8 and 11. The general findings are discussed in section 1.2.1 in the main part.

##### Full estimation: testing and the Easter break

Our preferred theoretical interpretation of the empirical findings in figure 1 results from calibrating one contact rate all throughout the OuS period and the Easter effect. The calibrated values are in line 3 “OuS & Easter throughout” in table 6. While the contact rate *a* does not differ strongly from the first line without the Easter break, the detection rate *λ*^test^ drops by around 50%. This is not surprising, however: When more individuals with symptoms do not go and visit a doctor (*x* is positive), the pool of non-reported individuals is larger. We therefore obtain a lower detection rate to match the same observed number of positive tests. The plot corresponding to line 3 is in figure 9. We see that the fit of cumulative cases (top left) and number of positive rapid tests (bottom left) are as good as they were with the earlier calibration visualized in figure 11.

**Figure 9:**
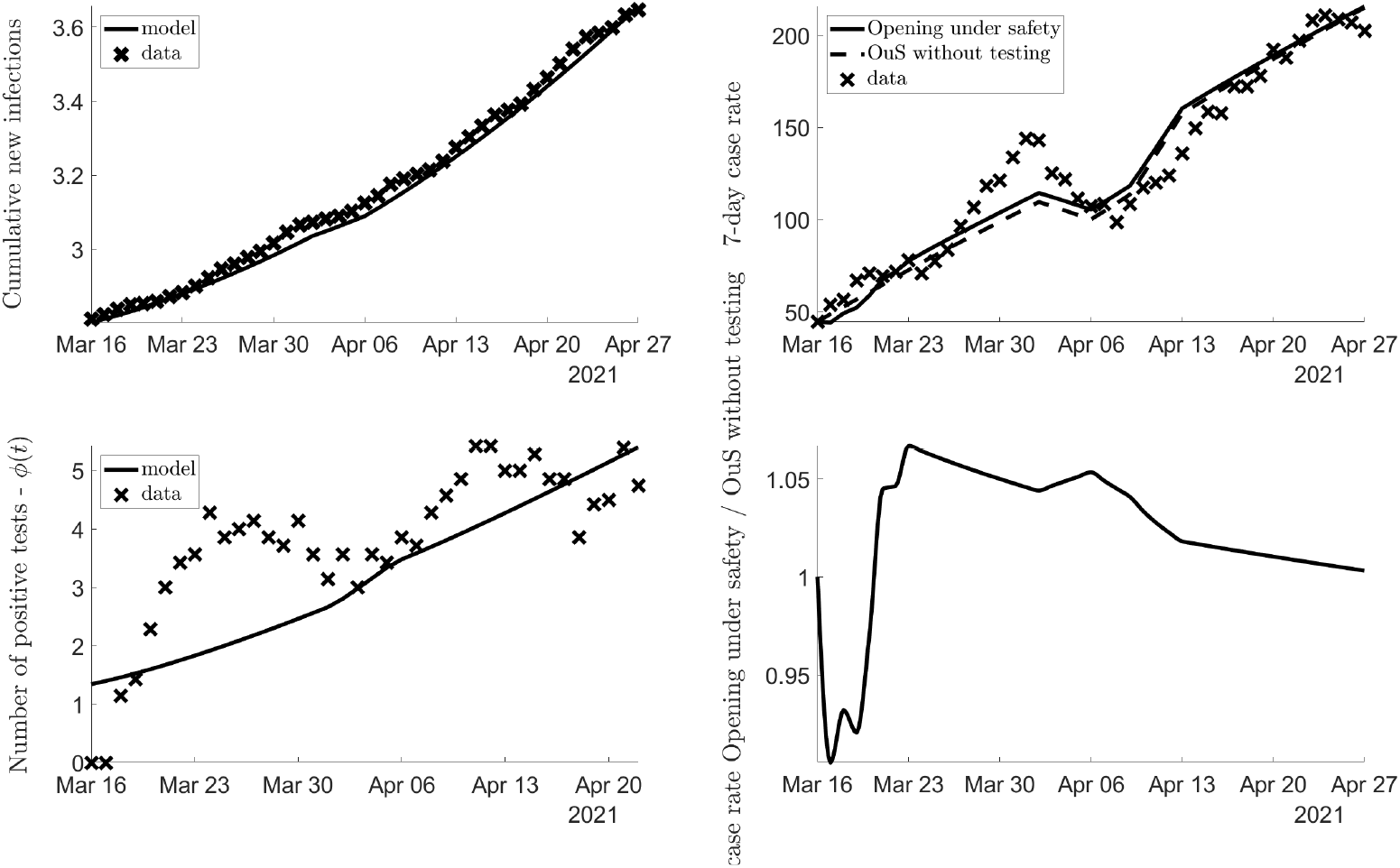
OuS and Easter break for identical parameters

Most importantly for our view, the downturn of the case rate during the Easter break can be easily explained by a reduction of testing and reporting. Our calibration shows that *x* = 33.4% of individuals do not go to emergency units during public holidays but rather stay at home. This leads to a temporary drop in case rates. (Conceptually, the drop is the same in the control region. In levels, case rates almost become identical.) After the Easter break, case rates increase again.

While the drop is smaller in the model than in the data, the model perfectly replicates the increase in case rates over the entire OuS period. Hence, we conclude that the permanent effect of OuS led to the rise in cases in Tübingen over time, the temporary dip is due to the Easter break.

When we quantify the testing effect plus the Easter Break and allow for ‘before Easter’ and ‘as of Easter effects’ on contact rates, we get the fourth and fifth line in table 6 for Tübingen. Note that it makes sense to distinguish these two sub-periods as visible in figure 3 (left). This figure clearly shows that the average number of visitors dropped to around 50% after Easter.

This is perfectly consistent with the drop in the contact rate *a* in line 4 ‘OuS before Easter’ to the value in line 5 ‘OuS as of Easter’.

The share of individuals not visiting a doctor is somewhat higher at 39.4% under this specification. The figure for this specification is almost indistinguishable from figure 9. We see this as further confirmation of our interpretation of the baseline findings of our SCM analysis.

A final thought experiment might further strengthen our point. Imagine, there is no testing at all during holidays. Let the OuS region have a high and the control region a low case rate just before holidays. Then, during holidays, case rates drop to zero. Regions seem to be identical in terms of reported cases. More generally speaking, the *relative* drop in case rates is the same across regions over holidays. As the difference is measured in *levels*, the difference falls.

In the background, however, the pandemic continues, it is simply not measured. When holidays are over, visits at GPs and testing resume and the difference between the treated and the control region is again visible in the data. The fundamental and crucial difference in the regions consists in the difference in the contact rates.

### A.5 Methods

#### A.5.1 The Synthetic control method

The synthetic control method (SCM) is by now a well established strategy to measure the *treatment effect* of specific policy measures (see Section A.3 for references). It has proven to be a useful tool in the context of the Covid-19 pandemic to study e.g. the effect of making face masks mandatory or to quantify the effect of lockdown measures (*5, 18*).

Here we provide the details regarding SCM that are relevant for our analysis. First, we set up the *donor pool* : it includes 400 Germany counties (“Landkreise” und “kreisfreie Städte”). 34 of these are located in BW and hence in the same state as Tübingen county. We exclude counties adjacent to Tübingen county (listed in appendix A.2.2) from the donor pool given a high likelihood of spillovers. We consider alternative donor pools in order to corroborate our results.

Second, we construct a *synthetic control unit* as a weighted average across the counties in the donor pool. Note that the number of counties with non-negligible weight is not restricted by our procedure and may vary across specifications. The weights are selected on the basis of a minimum distance approach.

Specifically, we target a set of *predictor variables* for Tübingen county in the pre-treatment period (that is, before March 16) in order to determine county weights. Predictors consist of pandemic measures and non-pandemic measures. The choice of predictor variables is driven (partly by their availability and) mainly by the desire to identify comparable counties based on fundamental determinants driving the outcome variable. The predictor set includes observations for the *outcome variable* (infection rate). In an ideal world, one would include those variables as predictors which determine the evolution of the pandemic in a county. As these ideal predictors are not available, regional characteristics and lagged outcome variables serve as proxies for the latent true variables. As pandemic-independent variables, we include both population as well as district characteristics in our predictor set. To assure that the control group consists of counties with a similar pandemic situation prior to the treatment, we also take pandemic-dependent variables into account. They include case rates prior to treatment and case rates of neighboring counties.

SCM chooses the weights on the counties in the donor pool by minimizing the ‘root mean square prediction error’ (RMSPE) which quantifies the distance of the (weighted sum of) comparison counties to Tübingen prior to treatment. Doing so, the control unit resembles Tübingen in terms of these variables as closely as possible. In this way, we ensure that pre-treatment differences in trends of the outcome variable are equalized. Table 5 lists all predictor variables. They include all socio-economic characteristics that are a) available at the county level and b) may matter for infection dynamics. In addition, we include weekly averages for infection rates in the six weeks prior to treatment.

Formally, let **x**_**1**_ denote the (*k* × 1) vector of predictor varibles in Tübingen and let **X**_**0**_ denote a (*k* × *n*) matrix with observations in the counties included in the donor pool consisting of *n* counties. Let **w** denote a (*n* × 1) vector of country weights *w*_*j*_, *j* = 1, …, *n*. Then, the control unit is defined by a **w**^*∗*^ which minimizes the mean squared error

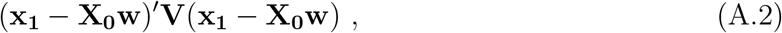

subject to *w*_*j*_ *>*= 0 for *j* = 1, …, *n* and 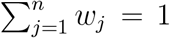. In this expression, **V** is a (*k* × *k*) symmetric and positive semidefinite matrix. Here, **V** is a weighting matrix assigning different relevance to the characteristics in **x**_**1**_ and **X**_**0**_. Although the matching approach is valid for any choice of **V**, it affects the weighted mean squared error of the estimator (*27*). We choose a diagonal **V** matrix such that the mean squared prediction error of the outcome variable (and the covariates) is minimized for the pre-treatment period (*27, 28*).

We conduct all SCM estimations in STATA using the SYNTH (*29*) and SYNTH RUNNER (*30*) packages. Our implementation follows largely (*5*).

Confidence intervals (CIs) are calculated from one-sided pseudo *p*-values obtained on the basis of comprehensive placebo-in-space tests. The latter tests calculate pseudo-treatment effects for all counties in the donor pool assuming that they, rather than Tübingen, were treated with OuS on March 16, 2021. We calculate one-sided pseudo *p*-values as the share of placebo-treatment effects that are larger than the observed treatment effects for treated counties. We thereby indicate the probability that the increase in the outcome variable was observed by chance given the distribution of pseudo-treatment effects in the donor pool.

To account for differences in pre-treatment match quality of the pseudo-treatment effects, only donors with a good fit in the pre-treatment period are considered for inference. Specifically, we do not include placebo effects in the pool for inference if the match quality of the control region, measured in terms of the pre-treatment root mean squared prediction error (RMSPE), is greater than 10 times the match quality of the treated unit (*31*). Based on the obtained pseudo *p*-values we calculate confidence intervals as described in (*32*).

#### A.5.2 A quantitative SIR model for Opening under Safety

To understand the effects of opening under safety (OuS), we start from an extended description of a pandemic illustrated in figure 10. Simpler models of this type have been employed e.g. by (*10*), (*33*) or (*34*).

**Figure 10:**
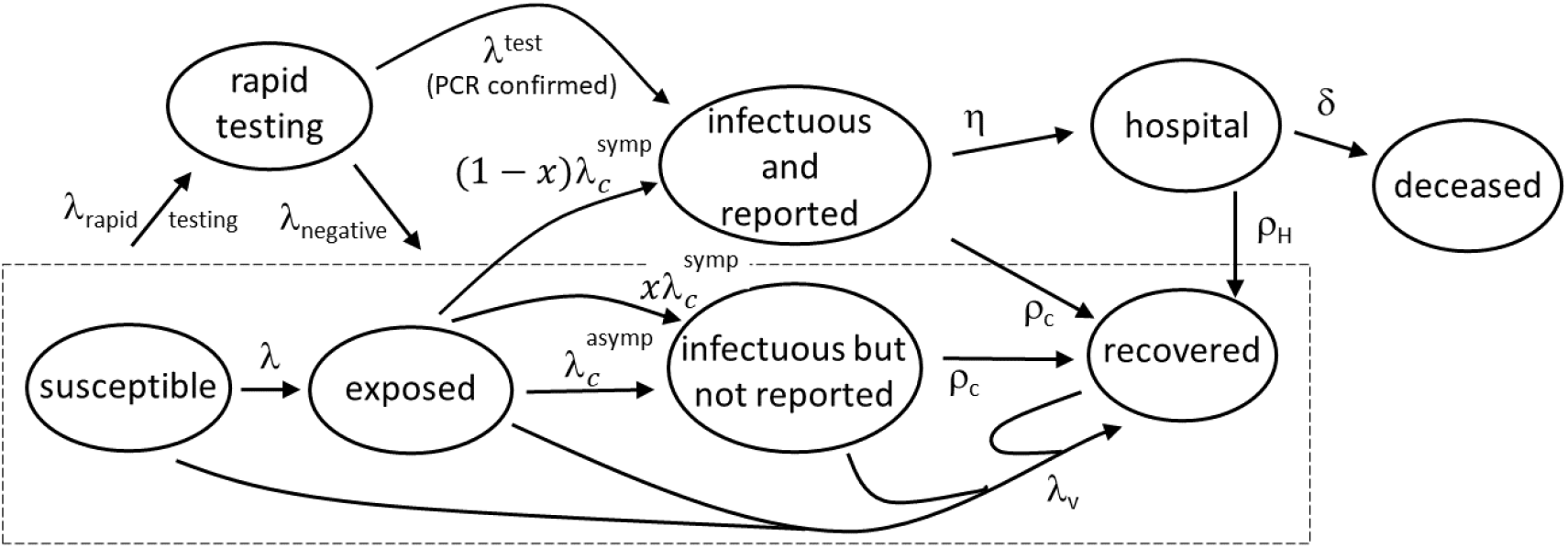
A SEIR model for opening under safety and the effect of rapid testing

##### A SIR model with testing and public holidays

Susceptible individuals *S* (*t*) can be infected after which they are exposed *E* (*t*). Exposed individuals can turn infectious and reported, *I*_*r*_ (*t*), or infectious but not reported, *I*_*n*_ (*t*). We assume that (‘standard’, i.e. non-rapid) tests are undertaken only for the *E* (*t*) - *I*_*r*_ (*t*)-flow, i.e. if individuals visit a doctor and display symptoms related to Covid-19. All reported infections are therefore symptomatic infections (Covid-19 cases). Tests employed in this case are PCR tests. Individuals can recover, *R* (*t*), or enter a hospital *H* (*t*). They can recover from hospital or die, *D* (*t*). The flows between the different states are illustrated in figure 10.

This figure extends earlier SIR models in two ways that are crucial to understanding OuS in Tübingen: the testing channel and public holidays. As the dashed rectangle is intended to show, rapid testing is applied to all individuals at rate *λ*^rapid testing^ that are not known to be infections (*I*_*r*_), in hospital (*H*) or dead (*D*). Some rapid tests are negative (at rate *λ*^negative^) and individuals stay in the state in which they were before being tested. For some individuals, however, rapid testing leads to a flow from asymptomatic, i.e. non-reported infectious individuals (*I*_*n*_), to reported infectious individuals (*I*_*r*_). Only non-reported infectious individuals can be detected to be infectious. This flow, visible via the rate *λ*^test^ in the figure and in equations (A.6) and (A.7) below, takes place only from *I*_*n*_ to reported infectious *I*_*r*_. We call it the detection rate. This flow is the big promise of OuS. Rapid negative tests do not imply any flows and are therefore not visible in the equations below. Only the net flow is visible in the equations as *λ*^test^ = *λ*^rapid testing^ - *λ*^negative^. The testing rate *λ*^rapid testing^ describes the rate with which OuS participants are tested in Tübingen. The rate *λ*^negative^ determines flows due to negative rapid tests and the detection rate *λ*^test^ is the residual.

The second extension captures the effect of public holidays at Easter on the number of reported infections. During public holidays, a share *x*(*t*) of individuals that feel sick stay at home. A share 1 — *x*(*t*) goes to emergency care. This is visible as 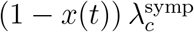 in (A.6) and as 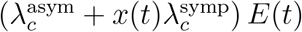 *E*(*t*) in (A.7) below. Figure 10 also displays this redirected flow 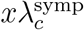. The idea behind *x*(*t*) is simply that some individuals with symptoms would go to a GP during workdays but do not do so during public holidays. When public holidays are over, symptoms are weak or gone, they also do not go to the GP on the next workday. A case that would have been (tested and) counted on workdays is not counted during holidays. To capture the effects of the Easter break from Friday to Monday, we set

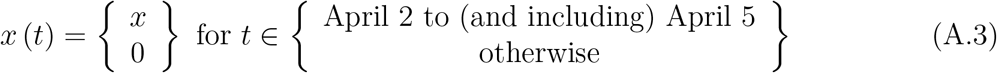

The differential equation system resulting from the underlying continuous time Markov chain (see (*35*) for more background) reads

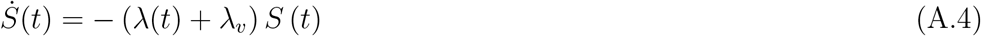

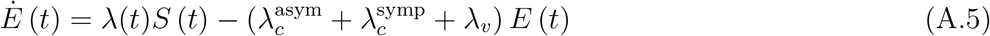

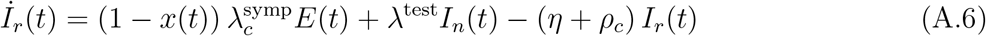

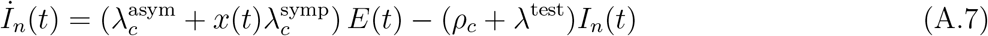

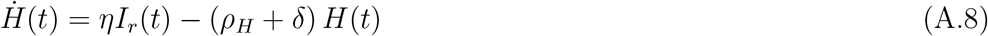

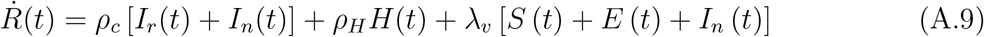

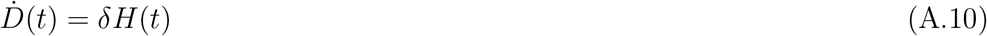

Some of our transition rates are functions of states of the system. The rate of getting infected is given by *λ*(*t*) = *ap* (*I*_*n*_) *π*, where *a* is the rate with which an individual meets other individuals, *p* (*I*_*n*_) is the probability that this individual gets in contact with a person being infectious and *π* is the probability that this leads to an own infection. In terms of an example, *a* is governed by how socially oriented a person is and by contact restrictions, *p* (*I*_*n*_) follows from the epidemic state and *π* is a function of the virus. As two examples, the UK-mutant or the delta-variant have a higher *π* than the original one.

The share of infectious individuals in all individuals one can meet in our model is given by *I*_*n*_*/* (*S* + *E* + *I* + *R*). In other words, we assume that there are no contacts with individuals in state *I*_*r*_ (they are in perfect quarantine), with individuals in hospital or with deceased individuals. Allowing for some non-linearity in arrival rates in the spirit of (*33*), we specify the infection rate as

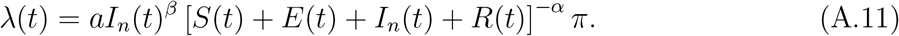

##### The effect of rapid testing

Now imagine the model is solved numerically. How do we study the effect of rapid testing on the pandemic? Note that the flow (not the rate) from *I*_*n*_ to *I*_*r*_ per unit of time is

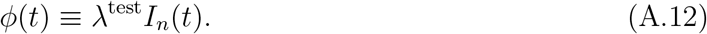

This flow is observed and illustrated in the right panel of figure 3. Data provides us with the number of (PCR confirmed) positive rapid tests per week or day (or any other unit of time). Let us call this number pos(*t*). If we choose one unit of time in our SIR model to equal one day, then we can equate *φ*(*t*) with pos(*t*) for any day *t*. Computing a model solution with *φ*(*t*) = 0 then shows, how the pandemic would have evolved in the absence of rapid testing. The difference to *φ*(*t*) =pos(*t*) provides a quantitative measure of the positive effects of rapid testing.

##### Calibrating the model

The calibration of the model follows earlier work (*10, 33*). We would like to match seven-day case rates, cumulative cases and the number of positive PCR-confirmed positive tests. For the initial values of *H* and *D* we use the reported data for Tübingen. The control group consists of weighted values according to Table 1. For the currently infected persons we used the number of reported infections over the last 18 days and distribute them evenly across the states *I*_*r*_ and *I*_*n*_. We approximate the initial value of *E* by a backward looking measure adding the number of reported infections over the last 4 days. The number of recovered people is given by subtracting all states from *N*. We normalize population size to 100 and express all numbers as shares. Table 7 presents the exact values we use.

**Table 7:** Initial conditions for the SEIR model in %

| Table 7: Initial conditions for the SEIR model in % |  |  |  |  |  |  |  |
| --- | --- | --- | --- | --- | --- | --- | --- |
| Region | $S(0)$ | $E(0)$ | $I_r(0)$ | $I_n(0)$ | $H(0)$ | $D(0)$ | $R(0)$ |
| Tübingen | 97.19 | 0.04 | 0.09 | 0.09 | 0.0425 | 0.16 | 2.39 |
| Control | 97.66 | 0.04 | 0.09 | 0.09 | 0.072 | 0.16 | 1.89 |

We calibrate the parameter vector *θ* ={*a, α, β, η, ρ*_*c*_, *ρ*_*H*_, *λ*^test^} by fixing some parameters according to values taken from the literature, and by choosing *θ* ={*a, α, β, λ*^test^, *x*} such as to minimize the Euclidean distance between the reported values and the corresponding predictions of the model. Formally,

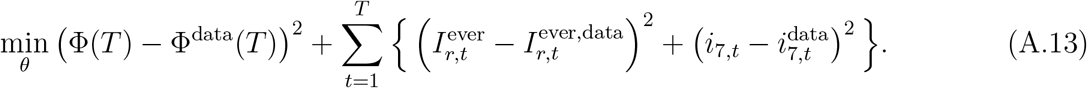

In terms of timing, *t* = 1 denotes March 16, 2021 and *t* = *T* is April 28, 2021. The number of positive (PCR confirmed) rapid tests at *T* is given by the integral over the flow, 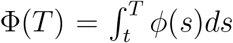. Similarly for the number of cumulative reported cases: Starting from our ODE system (A.4)-(A.10), numerical consistency suggests to add a differential equation which counts all reported cases 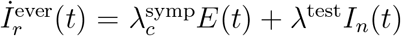. We solve this ODE jointly with the above system. Given the trajectory for 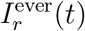,we can use it to calculate the 7-day case rate by differencing the time series:

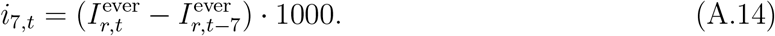

Since 7-day case rates are normalized per 100,000 inhabitants and we chose our population size as *N* = 100, we multiply the rate by 1,000 to scale the model rate accordingly to the reported rates. The 7-day case rate, 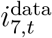 corresponds to the observed data.

We fix the remaining parameters by choosing *π* = 1 (which simply means that our estimate of the contact rate *a* is a joint estimate of *aπ*), a vaccination rate *λ*_*v*_ of 2% per week, and 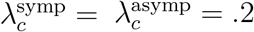, which corresponds to a mean incubation time of 5 days for both symptomatic and asymptomatic infections. We set the death rate to *δ* = .0268 and *η* = .1, *ρ*_*c*_ = 1*/*14, *ρ*_*h*_ = 1*/*21. Table 8 summarizes the fixed parameter values, table 6 presents the outcome of solving the minimization procedure in (A.13).

**Table 8:**
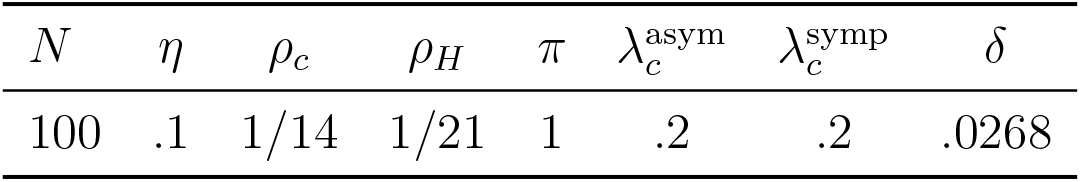
Fixed parameter values.

##### Calibration results

We start with a specification that ignores the Easter effect, i.e. for *x* = 0. Figure 11 provides an overview of the targeted data (cumulative cases, seven-day case rate and number of positive tests), the model fit and the ratio of case rates in the case of testing to the case of no testing. Calibrated parameter values for Tübingen and its control county are in line 1 (”OuS only”) and line 2 (”Control”) of table 6.

The top left panel of figure 11 shows cumulative new cases since 16th March, 2021. As the vertical axis shows, just below 2.8 percent of the population were infected on that date. The right panel shows the fit in terms of case rates. The crosses ‘data’ show the case rate as reported by health authorities. Comparing top left to top right shows that the model captures cumulative cases very well, but case rates to a lesser extent. The latter is not surprising as case rates or daily cases are much more volatile than cumulative cases. Yet, the overall tendency over the entire period of the experiment is matched perfectly for both variables. We can therefore confidently derive insights concerning the effect of testing from this calibration. Section A.4.3 will explain the volatility over Easter by reduced testing and reporting.

**Figure 11:**
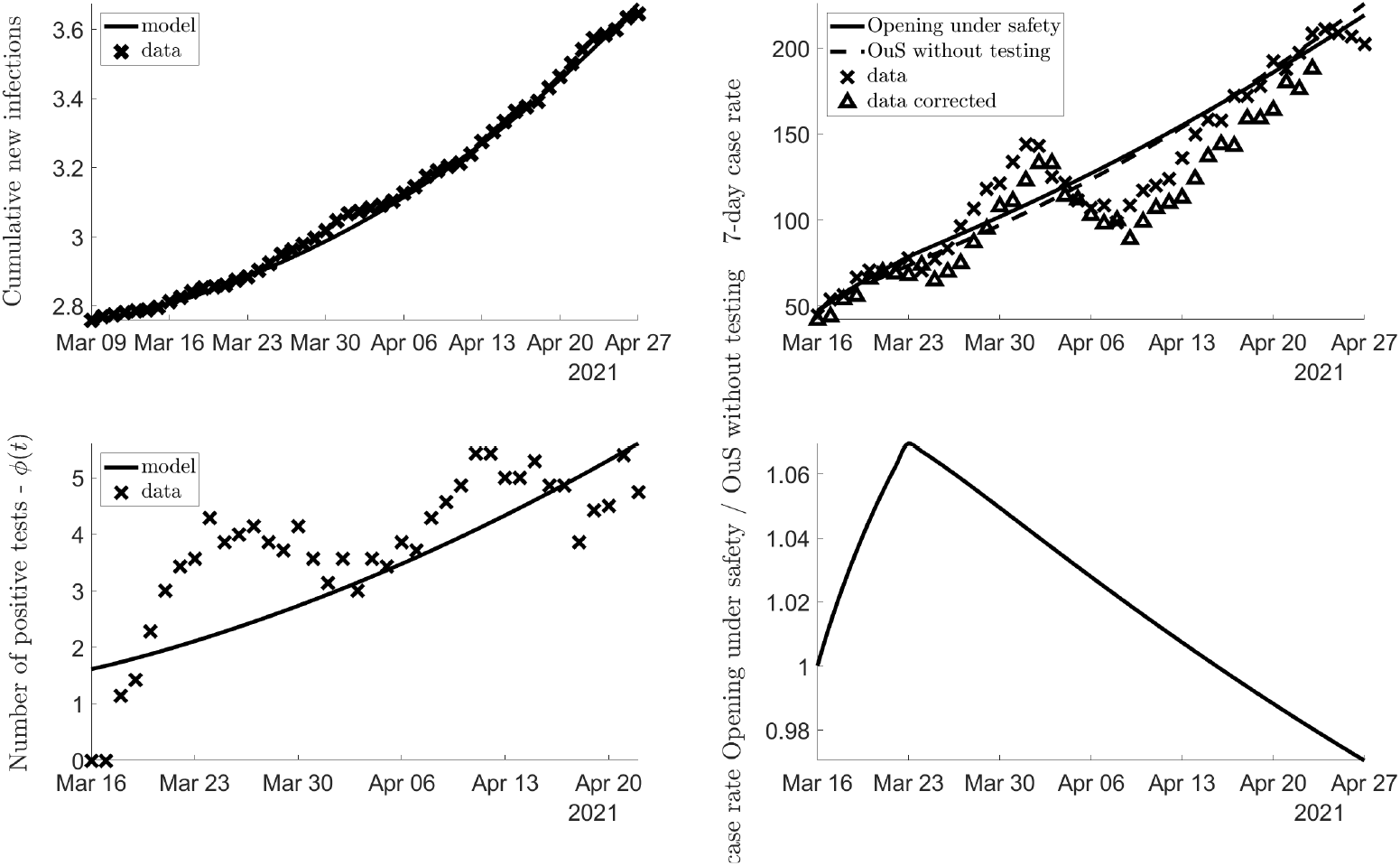
Data and paths of calibrated model and testing-no-testing ratio

The main purpose of employing this SIR variant consists in separating the effects of opening from the effects of testing. To obtain reliable results, the model needs to capture the total number of positive tests as well. This is achieved by estimating the detection rate *λ*^test^. The lower left panel shows that this also works sufficiently well. Again, the model does not capture short-run fluctuations. This, however, is of no importance for the overall trend and our overall finding. We referred to the lower right panel in section A.4.2.

### A.6 Discussion of donor pool

The a-priori choice of the set of counties from which SCM can choose comparison counties affects the composition of the synthetic twin of Tübingen. It is therefore crucial to understand the importance of the choice of the donor pool for the final result.

#### A.6.1 ‘Leave 1 out’

Having found our baseline specification for figure 1, we now proceed and check the robustness of the finding by evaluating the importance of individual counties in the synthetic twin of Tübingen. These counties are shown in table 1.

One standard approach to do this consists in removing each county shown in table 1, one by one, from the donor pool. Figure 12 shows the pre-treatment fit and the post-treatment effect of OuS under these adjusted donor pools. As the figure impressively shows, excluding any individual county from the donor pool does not affect our overall finding in any meaningful way. This robustness check strongly confirms our baseline finding.

**Figure 12:**
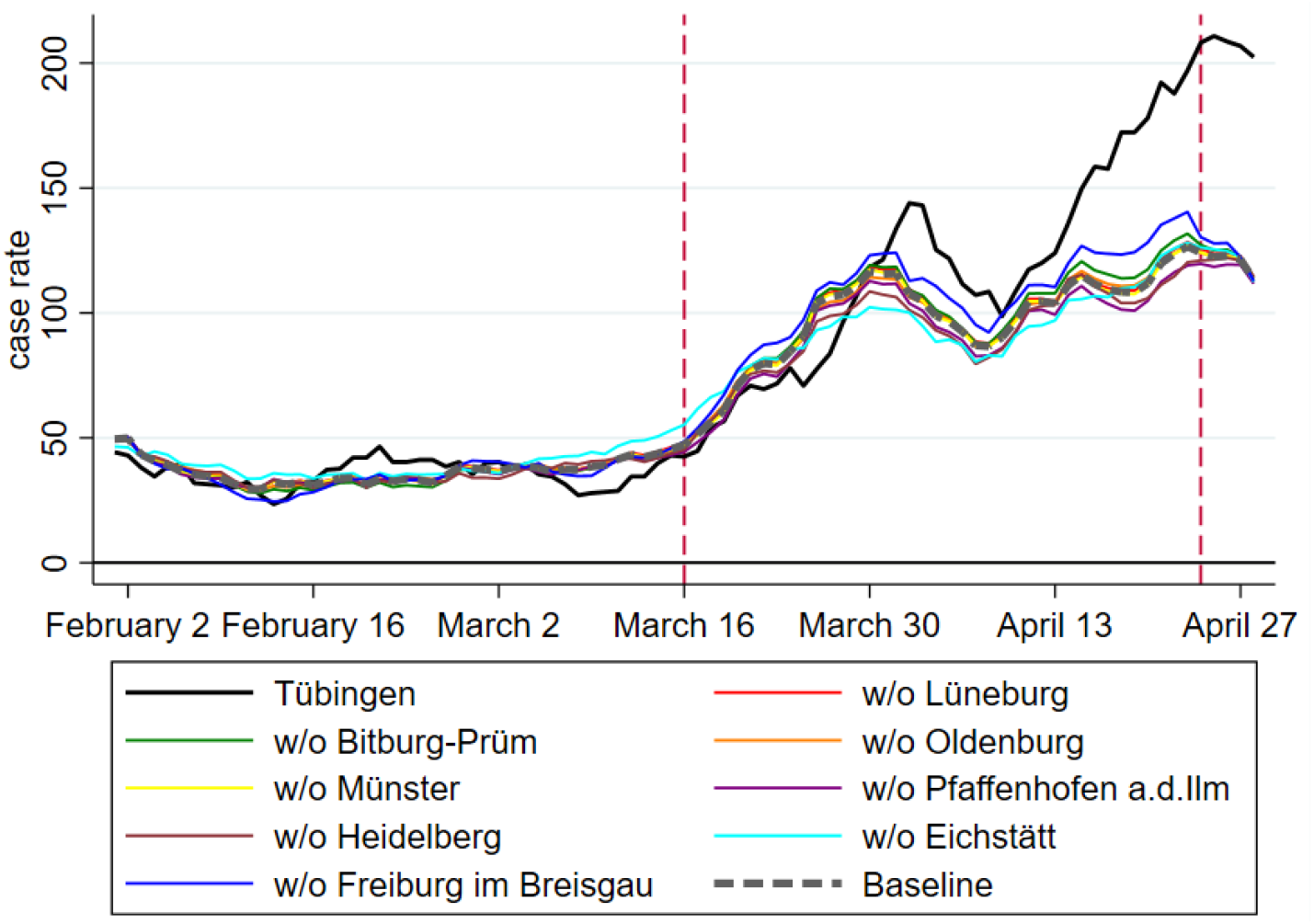
Leave one out for figure 15

#### A.6.2 Donor pool Baden-Württemberg only

The least restrictive donor pool consists of all German counties, excluding neighboring counties of Tübingen to avoid spillover effects. As can be seen in the left panel of figure 4, Tübingen county is at the lower end of the German case rate distribution before March 16. However, there are many other counties with case rates slightly above or below the case rate of Tübingen that can potentially serve as synthetic control group. For these reasons, our baseline specification works with this donor pool Germany.

The donor pool Baden-Württemberg (BW) is, by construction, more restrictive. The right panel of figure 4 shows the case rate of Tübingen county within the BW donor pool. Tübingen sometimes has the lowest case rate, especially shortly before the beginning of OuS on 16 March. Afterwards, it moved into the middle range. For these reasons, finding a suitable control group is difficult. This issue becomes even more prominent when discussing Tübingen city later on.

Nevertheless and for robustness checks, we now focus on a donor pool consisting of counties from Baden-Württemberg (BW) only. When we run SCM with a donor pool consisting of all BW counties (apart from neighbours), we obtain 5 control counties as seen in table 9. The corresponding predictor balance is in table 10 which also shows our predictor set ‘baseline’ in the first column.

**Table 9:** Control counties and their weights for figure 13

| Name | Weight |
| --- | --- |
| SK Heidelberg | 0.40 |
| LK Biberach | 0.24 |
| SK Freiburg i.Breisgau | 0.23 |
| LK Ostalbkreis | 0.11 |
| LK Tuttlingen | 0.01 |

Turning to results, the left panel of figure 13 shows the case rate in Tübingen (in black) and in the synthetic control county (dashed line). The right panel shows the difference between the case rates in Tübingen and in the control county. When we compare the overall outcome as visible in this figure with our baseline result in figure 1, we conclude that the difference between treatment and control is somewhat smaller in figure 13.

**Figure 13:**
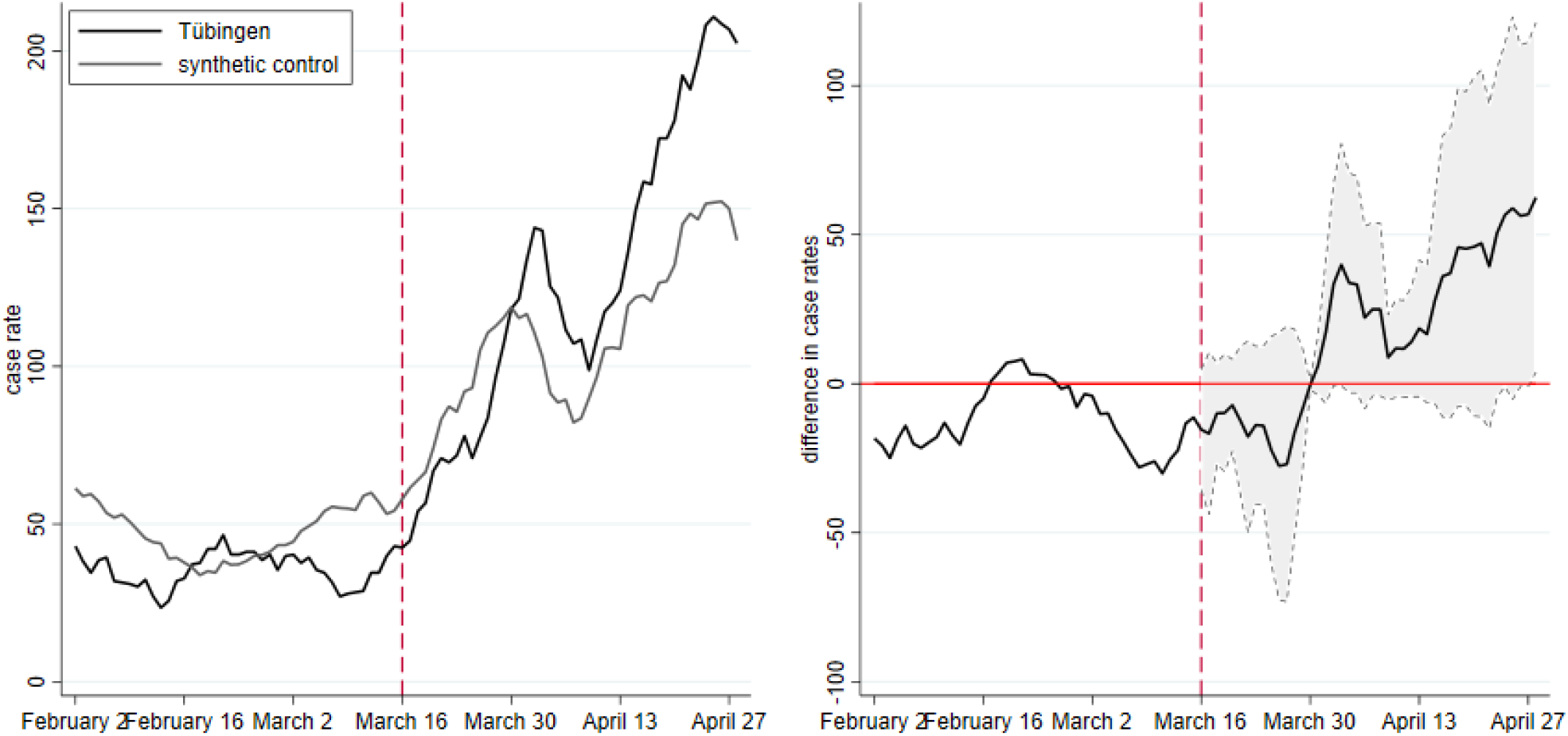
Case rates for donor pool Baden-Württemberg

When we look at the confidence intervall, we even see that the difference between Tübingen county and its synthetic twin from Baden-Württemberg is not significant. Nevertheless, the trend in the difference (right panel) is clearly positive and rising as of end of March. We do not put too much emphasis on this finding as the pre-treatment fit with donor pool Baden-Württemberg is not as good as in our baseline scenario. This finding therefore does not affect our overall conclusion, it rather confirms our baseline finding.

### A.7 Discussion of pandemic measure

An alternative measure of the pandemic state consists in normalized cumulative cases since, say, January 1st, 2021. Such a measure does not look at a (normalized) moving sum (like the seven-day case rate) but simply adds up the number of infections over time. It is therefore a measure with a much longer “memory”. An infection that occurred at any point since January 1st is always counted. Case rates “forget” cases that are older than, usually, 7 days. Cumulative cases are also normalized per 100K inhabitants to allow for a meaningful comparison across counties of different population size. We let then the SCM search for an appropriate comparison group for this dependent variable and compare the evolution of infections over time. Table 12 shows the fit between Tübingen and the synthetic twin city (table 11) for cumulative infections as dependent variable.

As the black and dashed curve before treatment on March 16 in figure 14 show, the fit between Tübingen and its synthetic twin county is almost perfect here. The better fit compared to case rates is not surprising as adding up infections since some starting date (1 January 2021 here) implies a smoother time series than adding infections over the previous 7 days. Table 12 shows the details of the fit.

**Figure 14:**
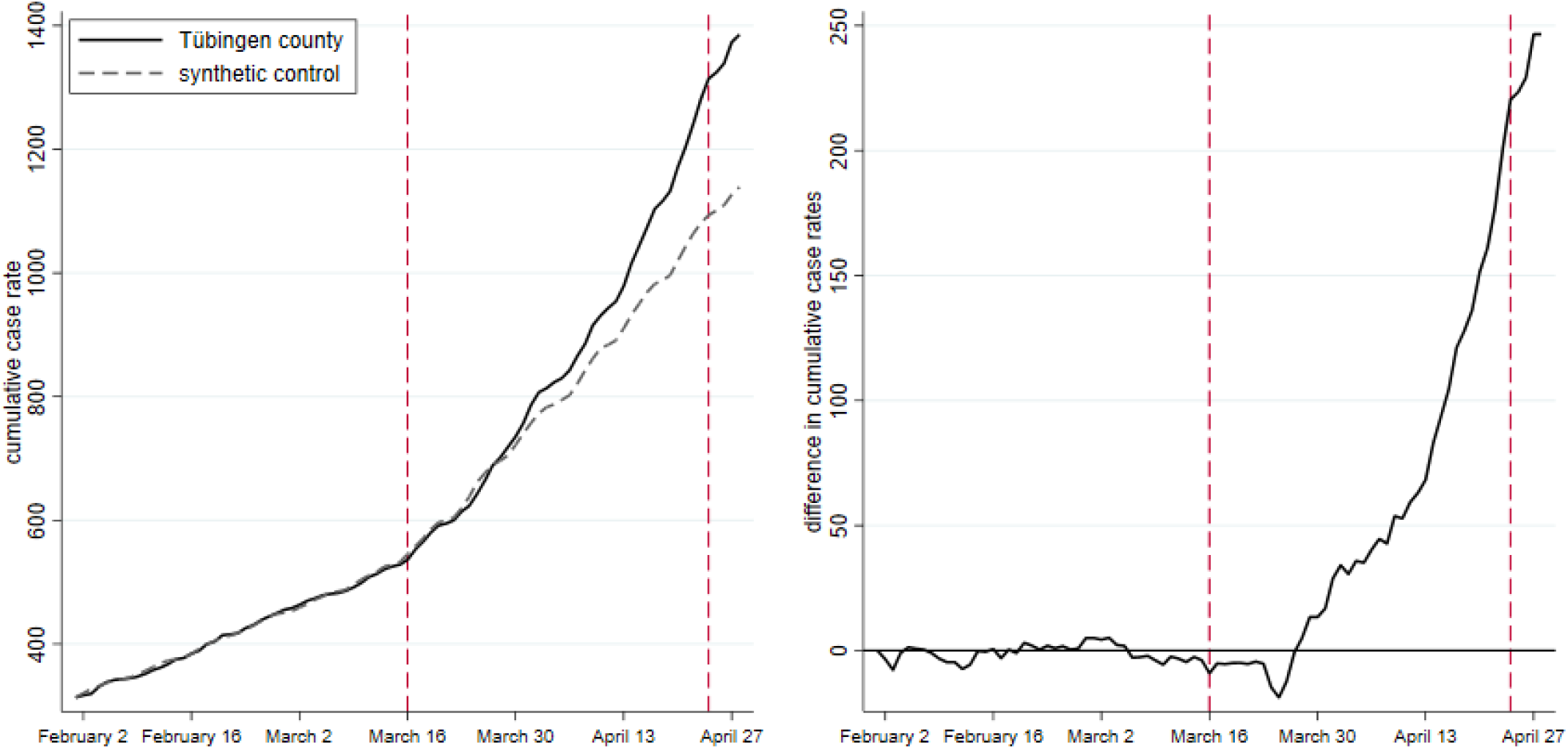
Cumulative cases per 100,000 since January 1st

What is much more important for our question, however, is the confirmation of our baseline result with this alternative measure. Tübingen county experiences an increase in cases relative to its synthetic control county.

### A.8 Discussion of predictor set

The predictor set is obviously important for the choice by SCM of control counties and their weights. Counties can be compared by fundamental characteristics like population density, educational background and medical services or by more pandemic-related features such as case rates prior to treatment.

As discussed in section 3.1, we have three basic predictor sets: predictor set ‘baseline’ (table 5), predictor set 1 (table 14) and predictor set 2 (table 16). Predictor set ‘baseline’ is also used for table 10 and table 12. In table 12, case rates are replaced by cumulative cases as our pandemic measure was changed to cumulative cases. A special predictor set ‘community’ is needed when we inquire into Tübingen city. See section A.9 for details.

**Table 10:** Balancing properties of predictor set ’baseline’ for figure 13

|  | Treated | Synthetic |
| --- | --- | --- |
| Seven-day case rate per 100k (Feb 1) | 44.30 | 60.78 |
| Seven-day case rate per 100k (Feb 8) | 31.45 | 53.33 |
| Seven-day case rate per 100k (Feb 15) | 31.89 | 39.25 |
| Seven-day case rate per 100k (Feb 22) | 40.31 | 37.12 |
| Seven-day case rate per 100k (Mar 1) | 39.87 | 44.11 |
| Seven-day case rate per 100k (Mar 8) | 27.02 | 55.17 |
| Seven-day case rate per 100k (Mar 15) | 42.97 | 54.93 |
| Cumulative cases over previous 7 days (Mar 9) | 63.00 | 110.45 |
| Cumulative cases over previous 14 days (Mar 15) | 158.00 | 223.74 |
| Average mobility (Mar 9 - Mar 15) | 0.00 | -0.09 |
| Average Temperature (Mar 9 - Mar 15) | 4.21 | 6.34 |
| Population density | 434.86 | 992.31 |
| Share of females in population | 51.26 | 51.15 |
| Average age of female population | 41.67 | 41.74 |
| Average age of male population | 40.03 | 39.77 |
| Old-age dependency ratio | 24.58 | 24.99 |
| Young-age dependency ratio | 20.20 | 19.33 |
| Medical doctors per population | 15.64 | 21.14 |
| Pharmacies per population | 23.48 | 27.61 |
| Categorical variable for population density of NUTS3 region | 2.00 | 1.61 |
| Share of highly educated persons in regional population | 26.47 | 29.29 |
| Stringency Index | 2.97 | 2.97 |
| Neighborhood (50km) seven-day case rate per 100k (Feb 1) | 81.64 | 89.76 |
| Neighborhood (50km) seven-day case rate per 100k (Feb 8) | 61.83 | 70.10 |
| Neighborhood (50km) seven-day case rate per 100k (Feb 15) | 51.14 | 51.46 |
| Neighborhood (50km) seven-day case rate per 100k (Feb 22) | 44.51 | 48.70 |
| Neighborhood (50km) seven-day case rate per 100k (Mar 1) | 58.36 | 51.36 |
| Neighborhood (50km) seven-day case rate per 100k (Mar 8) | 56.70 | 61.76 |
| Neighborhood (50km) seven-day case rate per 100k (Mar 15) | 70.73 | 77.87 |
| RMSPE (pre-treatment) | 19.57 |  |

*Note:* Dates in parentheses indicate when the respective variable was measured.

**Table 11:** Control counties and their weights for figure 14

| Name | Weight |
| --- | --- |
| LK Eichstätt | 0.19 |
| SK Münster | 0.18 |
| SK Freiburg i.Breisgau | 0.15 |
| SK Heidelberg | 0.13 |
| LK Lüneburg | 0.12 |
| LK Bitburg-Prüm | 0.01 |
| SK Trier | 0.06 |
| SK Rostock | 0.04 |
| SK Oldenburg | 0.04 |

**Table 12:** Balancing properties of predictor set ’baseline’ for figure 14

|  | Treated | Synthetic |
| --- | --- | --- |
| Cumulative cases per 100k (Feb 1) | 313.61 | 313.68 |
| Cumulative cases per 100k (Feb 8) | 344.62 | 345.59 |
| Cumulative cases per 100k (Feb 15) | 377.40 | 377.87 |
| Cumulative cases per 100k (Feb 22) | 417.71 | 417.36 |
| Cumulative cases per 100k (Mar 1) | 458.02 | 452.98 |
| Cumulative cases per 100k (Mar 8) | 485.93 | 488.05 |
| Cumulative cases per 100k (Mar 15) | 528.45 | 532.28 |
| Seven-day case rate per 100k (Mar 15) | 42.97 | 42.91 |
| Average mobility (March 9 - March 15) | 0.00 | -0.17 |
| Average Temperature (March 9 - March 15) | 4.21 | 5.54 |
| Population density | 434.86 | 798.44 |
| Share of females in population | 51.26 | 51.05 |
| Average age of female population | 41.67 | 42.49 |
| Average age of male population | 40.03 | 40.37 |
| Old-age dependency ratio | 24.58 | 26.19 |
| Young-age dependency ratio | 20.20 | 19.56 |
| Medical doctors per population | 15.64 | 18.60 |
| Pharmacies per population | 23.48 | 27.48 |
| Categorical variable for population density of NUTS3 region | 2.00 | 2.04 |
| Share of highly educated persons in regional population | 26.47 | 23.83 |
| Stringency Index | 2.97 | 2.90 |
| Neighborhood (50km) seven-day case rate per 100k (Feb 1) | 81.64 | 79.27 |
| Neighborhood (50km) seven-day case rate per 100k (Feb 8) | 61.83 | 64.92 |
| Neighborhood (50km) seven-day case rate per 100k (Feb 15) | 51.14 | 52.00 |
| Neighborhood (50km) seven-day case rate per 100k (Feb 22) | 44.51 | 54.04 |
| Neighborhood (50km) seven-day case rate per 100k (Mar 1) | 58.36 | 53.47 |
| Neighborhood (50km) seven-day case rate per 100k (Mar 8) | 56.70 | 53.14 |
| Neighborhood (50km) seven-day case rate per 100k (Mar 15) | 70.73 | 69.14 |
| RMSPE (pre-treatment) | 4.96 |  |

*Note:* Dates in parentheses indicate when the respective variable was measured.

**Table 13:** Control counties and weights for figure 15

| Name | Weight |
| --- | --- |
| SK Heidelberg | 0.45 |
| LK Eichstätt | 0.35 |
| SK Trier | 0.06 |
| SK Leipzig | 0.06 |
| LK Nordfriesland | 0.04 |
| LK Emsland | 0.04 |

**Table 14:**
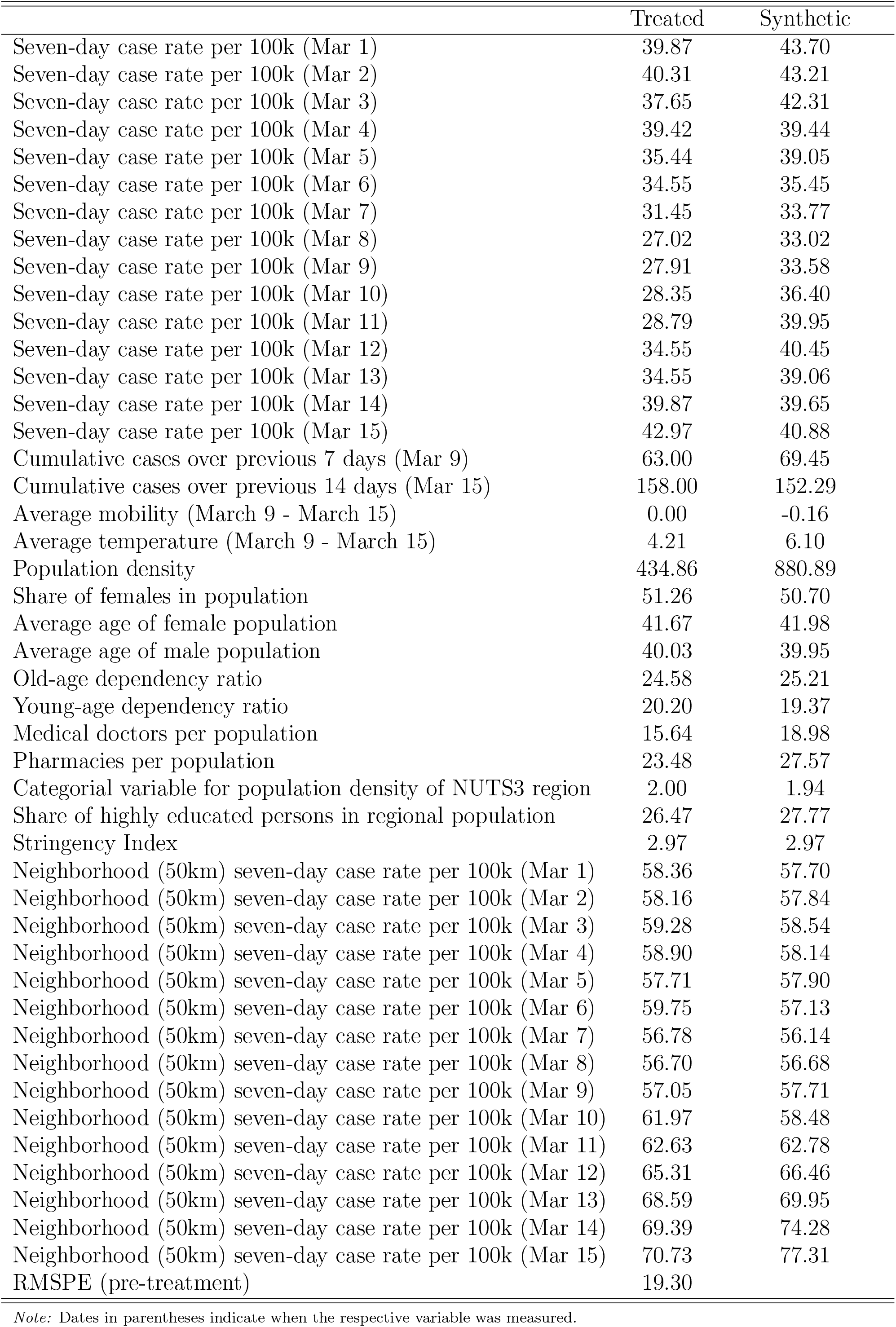
Balancing properties of predictor set 1 for figure 15

Our baseline predictor set includes the pandemic measure of Tübingen county and its neighbours over a period of six weeks (i.e. February 1 to March 15 in table 5) to ensure that the pandemic measure in the control group evolved in a similar way to the pandemic measure in Tübingen. We chose the matching period to be as long as the prediction period. One could argue however, that a good match on the pandemic measure dating back multiple weeks has no predictive power for the development of the pandemic and that a the pandemic measure should be included at a higher frequency to ensure a very good pre treatment fit. This idea is captured by predictor set 1 and 2 and visible e.g. in table 14 with daily case rates from March 1 to March 15.

#### A.8.1 Full donor pool and predictor set 1

We now display the results shown in our central robustness figure 2 in the main part in more detail. As figure 15 shows, including pre-treatment measures of the pandemic at higher frequency does not change the basic finding, even though the members of the synthetic control county change.

**Figure 15:**
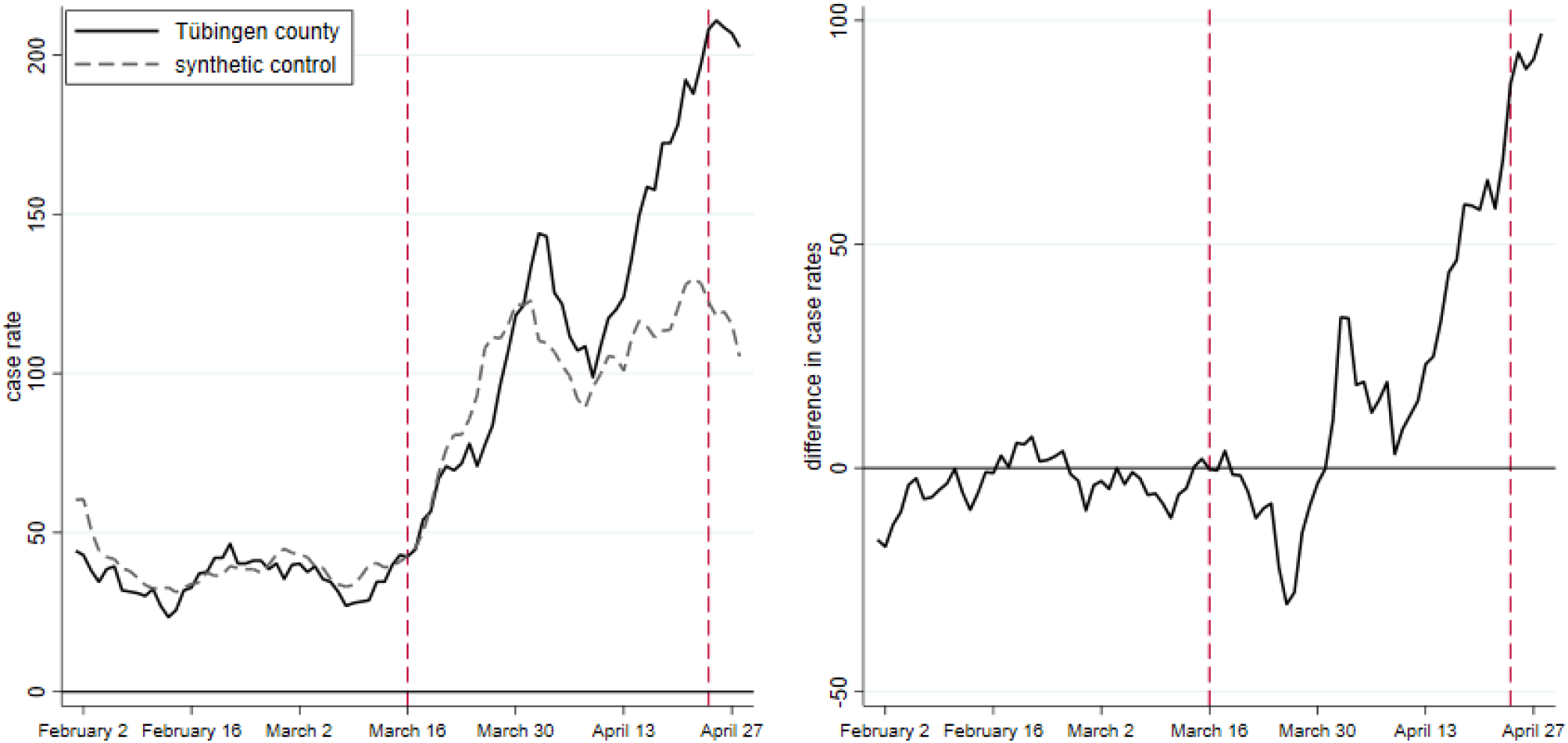
Seven-day case rate with predictor set 1

#### A.8.2 Full donor pool and predictor set 2

We now consider a further alternative to the baseline predictor set employed for figure 1. Our predictor set 2 is shown in table 16 and puts less weight on case rates in the matching period. Not surprisingly, we get a different set of comparison counties in table 15 and a modified predictor balance (table 16). The result is in figure 16.

**Table 15:** Control counties and their weights for figure 16

| Name | Weight |
| --- | --- |
| LK Eichstätt | 0.45 |
| SK Heidelberg | 0.31 |
| SK Freiburg i.Breisgau | 0.17 |
| LK Tuttlingen | 0.07 |
| SK Berlin | 0.003 |

**Table 16:** Balancing properties of predictor set 2 for figure 16

|  | Treated | Synthetic |
| --- | --- | --- |
| Seven-day case rate per 100k (Mar 1) | 39.87 | 40.84 |
| Seven-day case rate per 100k (Mar 8) | 27.91 | 35.40 |
| Seven-day case rate per 100k (Mar 15) | 42.97 | 39.98 |
| Cumulative cases over previous 7 days (Mar 9) | 63.00 | 65.79 |
| Cumulative cases over previous 14 days (Mar 15) | 158.00 | 138.40 |
| Average mobility (March 9 - March 15) | 0.00 | -0.13 |
| Average temperature (March 9 - March 15) | 4.21 | 5.64 |
| Population density | 434.86 | 783.67 |
| Share of females in population | 51.26 | 50.57 |
| Average age of female population | 41.67 | 41.71 |
| Average age of male population | 40.03 | 39.83 |
| Old-age dependency ratio | 24.58 | 24.67 |
| Young-age dependency ratio | 20.20 | 20.25 |
| Medical doctors per population | 15.64 | 18.22 |
| Pharmacies per population | 23.48 | 25.90 |
| Categorical variable for population density of NUTS3 region | 2.00 | 1.97 |
| Share of highly educated persons in regional population | 26.47 | 26.09 |
| Stringency index | 2.97 | 2.99 |
| Neighborhood (50km) seven-day case rate per 100k (Mar 1) | 58.36 | 47.05 |
| Neighborhood (50km) seven-day case rate per 100k (Mar 8) | 57.05 | 52.26 |
| Neighborhood (50km) seven-day case rate per 100k (Mar 15) | 70.73 | 72.11 |
| RMSPE (pre-treatment) | 20.00 |  |

*Note:* Dates in parentheses indicate when the respective variable was measured.

**Figure 16:**
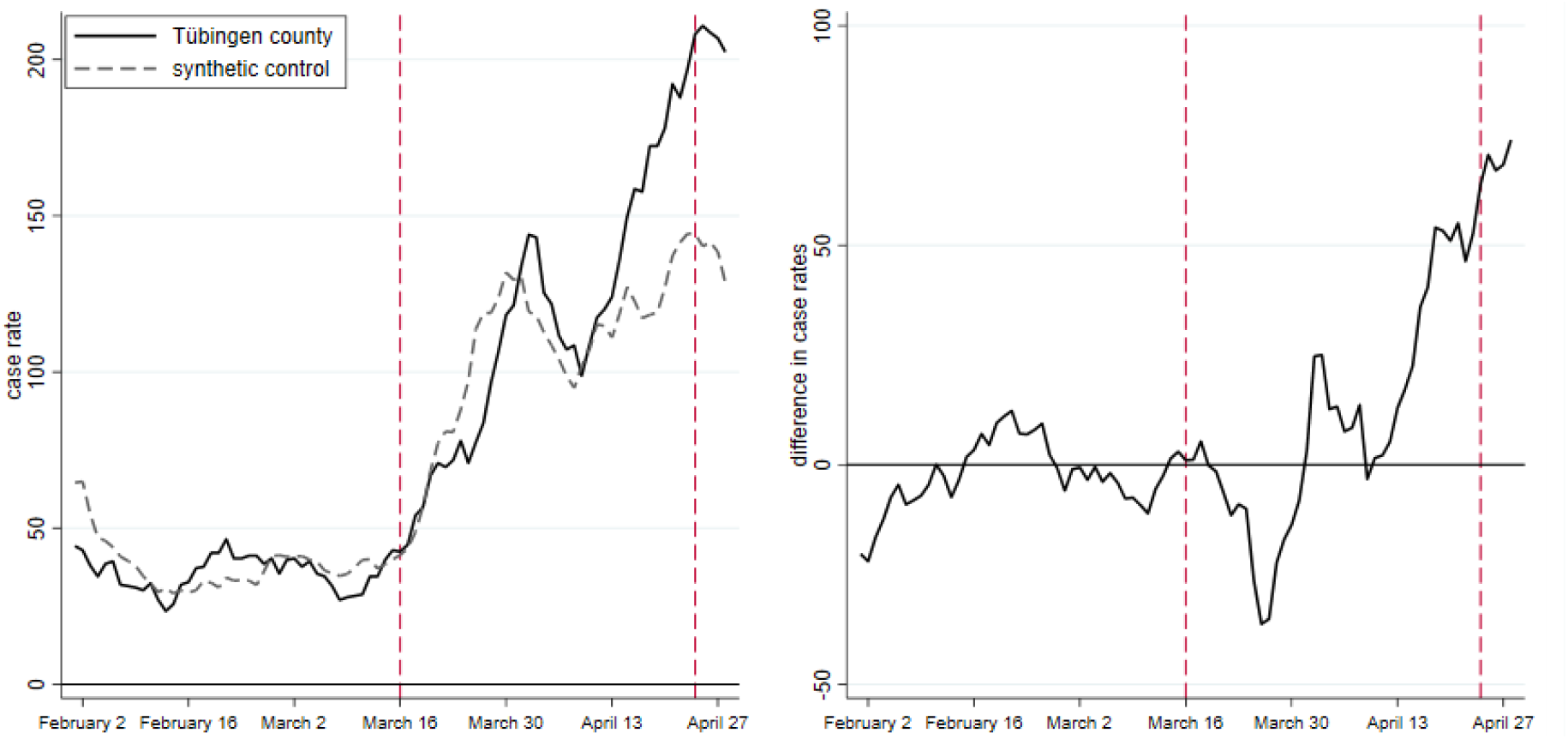
Seven-day case rates for predictor set 2

As in figure 1, the left panel in figure 16 shows case rates in Tübingen (solid) and in the synthetic control county (dashed). The right panel shows their difference. As for predictor set 1, we get similar overall results. Again, our baseline specification is confirmed.

#### A.8.3 The effect of spatial controls

One potential alternative explanation of why we see an increase in reported infection rates in Tübingen but only to a much smaller extent in the synthetic control could be that the pandemic development in neighboring counties affects the development in Tübingen. If, e.g., Tübingen before the beginning of OuS was surrounded by counties with high case rates, it is possible that these infections may spill over to Tübingen county. One key mechanism through which this may happen in the absence of OuS is through commuters. If that was the case, we would potentially observe an increase in case rates in Tübingen that is not due to OuS, but rather due to Tübingen’s proximity to regions with high infection rates.

To check for this potential explanation, our baseline regression matches Tübingen with potential control regions that exhibit similar case rates in their respective neighboring counties. To this end, we construct a new, additional set of predictors, to which we refer as spatial predictors. For every week in our pre-treatment period and for every county in Germany, we calculate the average infection rates (reported infections over 7 days per 100,000 inhabitants) across neighboring counties. We define neighboring counties as all counties whose centroids are not more than 50 km away from the focal county’s centroid. Using other definitions for example adjacent neighbors or a 90 km centroid definition did not change our results.

Our robustness check in figure 2 shows, again, that the main result does not change. We therefore conclude that the increases in cases in Tübingen relative to its synthetic twin is not driven by infection rates in neighbouring counties creating a surge in cases in Tübingen.

### A.9 Does Tübingen city differ from Tübingen county?

The final check of our findings consists in treating the *city* of Tübingen as an independent unit. The county of Tübingen is then also excluded from the donor pool, as are neighboring regions of the county of Tübingen (as in all other donor pools). Figure 17 provides some geographical background.

**Figure 17:**
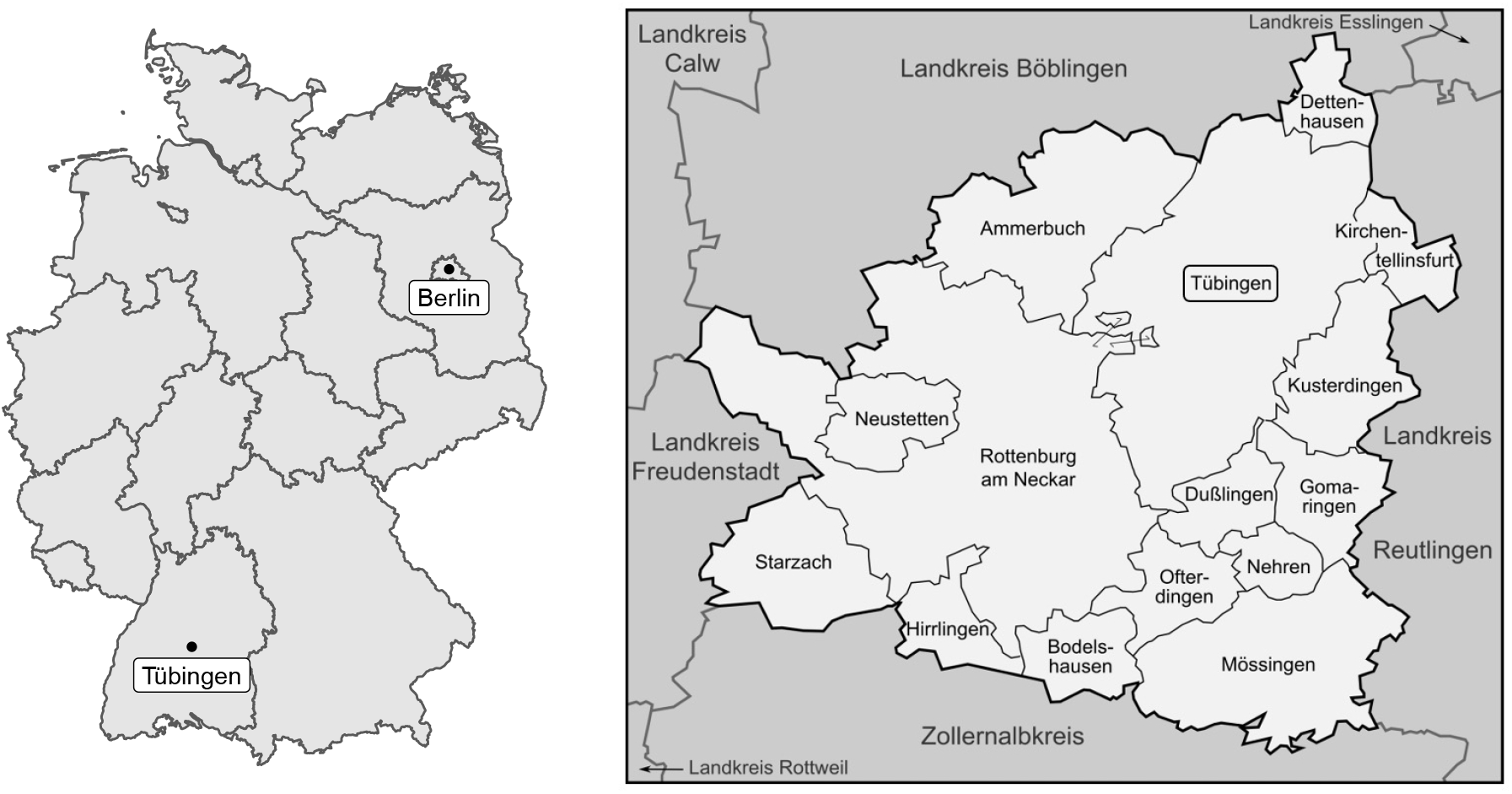
The position of Tübingen county within Germany and its communities. Left map copyright GeoBasis-DE / BKG (2021), right map sourced from (*36*).

The effect of OuS on Tübingen city can be understood only with an adjusted predictor set. Data used in our predictor sets used so far are not all available for Tübingen city. We therefore collected information on Tübingen city and grouped all available variables in our new predictor set ‘community’ shown in table 18. Variables found for Tübingen city were then also used for all other counties in Germany.

#### A.9.1 The effect of the predictor set community for Tübingen county

Imagine we find some result on Tübingen city that differs from our baseline finding. Given that we changed both the region of analysis and the predictor set, we would not know whether the differential effect is due to our focus on Tübingen city. It could be due to the new predictor set ‘community’.

To rule this out, we now replicate our baseline analysis with the predictor set ‘community’ instead of the predictor set ‘baseline’. The resulting synthetic control group is described in table 17.

**Table 17:** Weights for figure 18 (left)

| name | weight |
| --- | --- |
| LK Eichstätt | 0.38 |
| LK Lüneburg | 0.20 |
| SK Oldenburg | 0.12 |
| SK Brandenburg a.d.Havel | 0.09 |
| SK Rostock | 0.07 |
| SK Heidelberg | 0.05 |
| SK Neustadt a.d.Weinstraße | 0.02 |
| SK Frankenthal | 0.02 |
| SK Erlangen | 0.02 |
| LK Ebersberg | 0.01 |
| SK Osnabrück | 0.01 |
| LK Steinburg | 0.01 |
| LK Kusel | 0.01 |

**Table 18:** Balancing properties of predictor set community for figure 18

|  | Treated | Synthetic |
| --- | --- | --- |
| Seven-day case rate per 100k (Feb 9) | 31.01 | 31.25 |
| Seven-day case rate per 100k (Feb 15) | 31.89 | 31.98 |
| Seven-day case rate per 100k (Feb 22) | 40.31 | 40.21 |
| Seven-day case rate per 100k (Mar 1) | 39.87 | 39.91 |
| Seven-day case rate per 100k (Mar 8) | 27.02 | 27.53 |
| Seven-day case rate per 100k (Mar 15) | 42.97 | 42.77 |
| Cumulative cases over previous 7 days (Mar 15) | 97.00 | 59.38 |
| Population density | 434.86 | 516.46 |
| Share of females in population | 51.26 | 50.50 |
| Young-age dependency ratio | 20.20 | 20.94 |
| Old-age dependency ratio | 24.58 | 29.79 |
| Incommuting | 64.86 | 64.62 |
| Outcommuting | 67.73 | 63.90 |
| Accessibility | 13.00 | 15.56 |
| Pharmacies per population | 23.48 | 24.25 |
| Transport | 99.00 | 89.71 |
| Stringency index | 2.91 | 2.89 |
| Neighborhood (50km) seven-day case rate per 100k (Feb 8) | 61.62 | 61.52 |
| Neighborhood (50km) seven-day case rate per 100k (Feb 15) | 51.14 | 52.02 |
| Neighborhood (50km) seven-day case rate per 100k (Feb 22) | 44.51 | 51.10 |
| Neighborhood (50km) seven-day case rate per 100k (Mar 1) | 58.36 | 54.75 |
| Neighborhood (50km) seven-day case rate per 100k (Mar 8) | 56.70 | 54.28 |
| Neighborhood (50km) seven-day case rate per 100k (Mar 15) | 70.73 | 70.75 |
| RMSPE (pre-treatment) | 3.19 |  |

*Note:* Dates in parentheses indicate when the respective variable was measured.

**Table 19:** Weights for figure 19 left and right

| name | weight | name | weight |
| --- | --- | --- | --- |
| LK Eichstätt | 0.23 | LK Enzkreis | 0.41 |
| SK Osnabrück | 0.16 | SK Ulm | 0.17 |
| LK Dachau | 0.16 | LK Konstanz | 0.13 |
| LK Lüneburg | 0.10 | SK Heidelberg | 0.13 |
| SK Brandenburg a.d.Havel | 0.08 | LK Rhein-Neckar-Kreis | 0.11 |
| SK Oldenburg | 0.08 | LK Heilbronn | 0.05 |
| LK Konstanz | 0.07 | LK Rottweil | 0.001 |
| LK Ebersberg | 0.06 |  |  |
| SK Landshut | 0.02 |  |  |
| LK Plön | 0.02 |  |  |
| SK Bottrop | 0.01 |  |  |

**Table 20:** Balancing properties of predictor set community for figure 20

|  | Treated | Synthetic |
| --- | --- | --- |
| Seven-day case rate per 100k (Feb 8) | 0.31 | 0.32 |
| Seven-day case rate per 100k (Feb 15) | 0.81 | 0.81 |
| Seven-day case rate per 100k (Feb 22) | 0.50 | 0.50 |
| Seven-day case rate per 100k (Mar 1) | 0.63 | 0.61 |
| Seven-day case rate per 100k (Mar 8) | 0.31 | 0.32 |
| Seven-day case rate per 100k (Mar 15) | 0.19 | 0.07 |
| Cumulative cases over previous 7 days (March 15) | 19.00 | 53.64 |
| Population density | 827.00 | 733.88 |
| Share of females in population | 52.50 | 51.00 |
| Young-age dependency ratio | 16.90 | 18.83 |
| Old-age dependency ratio | 20.40 | 35.53 |
| Incommuting | 62.10 | 61.61 |
| Outcommuting | 43.40 | 51.13 |
| Accessibility | 0.00 | 21.79 |
| Pharmacies per population | 80.00 | 34.65 |
| transport | 100.00 | 95.45 |
| Stringency index | 2.91 | 2.80 |
| Neighborhood (50km) seven-day case rate per 100k (Feb 9) | 61.62 | 68.62 |
| Neighborhood (50km) seven-day case rate per 100k (Feb 15) | 51.14 | 56.46 |
| Neighborhood (50km) seven-day case rate per 100k (Feb 22) | 44.51 | 58.10 |
| Neighborhood (50km) seven-day case rate per 100k (Mar 1) | 58.36 | 58.73 |
| Neighborhood (50km) seven-day case rate per 100k (Mar 8) | 56.70 | 59.86 |
| Neighborhood (50km) seven-day case rate per 100k (Mar 15) | 70.73 | 73.99 |
| RMSPE (pre-treatment) | 0.27 |  |

*Note:* Dates in parentheses indicate when the respective variable was measured.

**Table 21:**
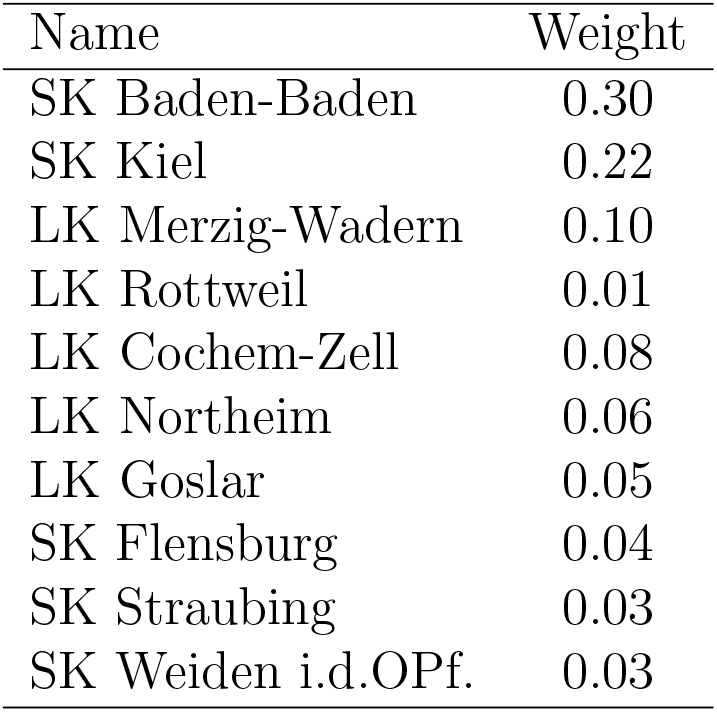
Weights for figure 20

The left part of figure 18 shows us the effect of changing the predictor set from ‘baseline’ to ‘community’. The right panel replicates our baseline figure 1. As we see, the effect of OuS under the predictor set ‘community’ appears somewhat larger than in the baseline scenario. Practically speaking, however, the findings are the same. Hence, any change we might find in what follows for the city of Tübingen as compared to Tübingen county cannot be due to the use of the new predictor set ‘community’.

**Figure 18:**
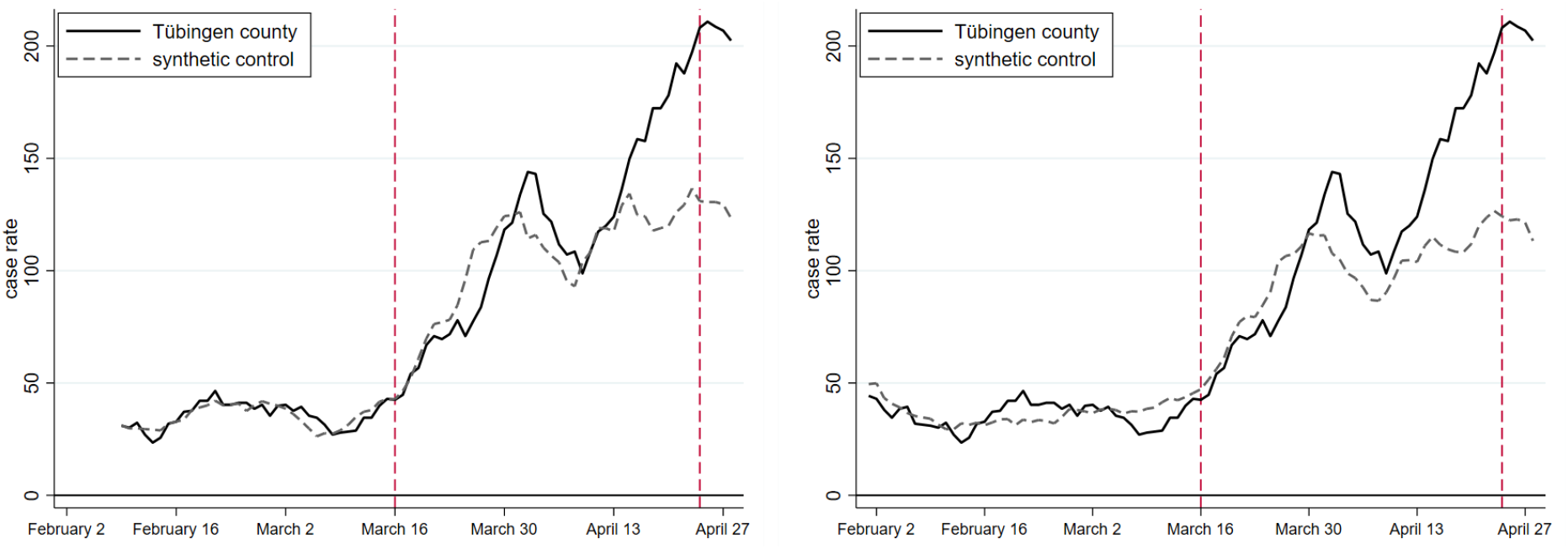
Tübingen county with predictor set ’community’ (left) and baseline (right)

#### A.9.2 Studying growth rates: Normalization

Finding control counties for Tübingen city with seven-day case rates as the pandemic measure turned out to be very difficult. This is due to the fact that Tübingen city had very low case rates (as shown earlier in figure 4). The pre-treatment fit and the predictor balance were very bad in an initial SCM analysis. This might also be due to the fact that SCM allows for positive weights only. We therefore normalized seven-day case rates such that they equal zero on March 16 for all counties (and for Tübingen city, as also shown earlier in figure 5).

##### Tübingen county

We now re-perform our baseline SCM with these normalized case rates. Figures then display growth factors of cases rates (or growth rates if normalized by 100). Figure 19 plots time on the horizontal and growth factors on the vertical axis. It shows that case rates in Tübingen county increased by a factor of 4 as compared to an increase in case rates in the synthetic control county by a factor of 2. When we restrict the donor pool to Baden-Württemberg in the right panel, the same ratios can be found. Hence, working with growth rates of case rates as opposed to case rates in our baseline analysis leads to the same overall conclusion that we stress in the main text. This allows us to finally study the city of Tübingen.

**Figure 19:**
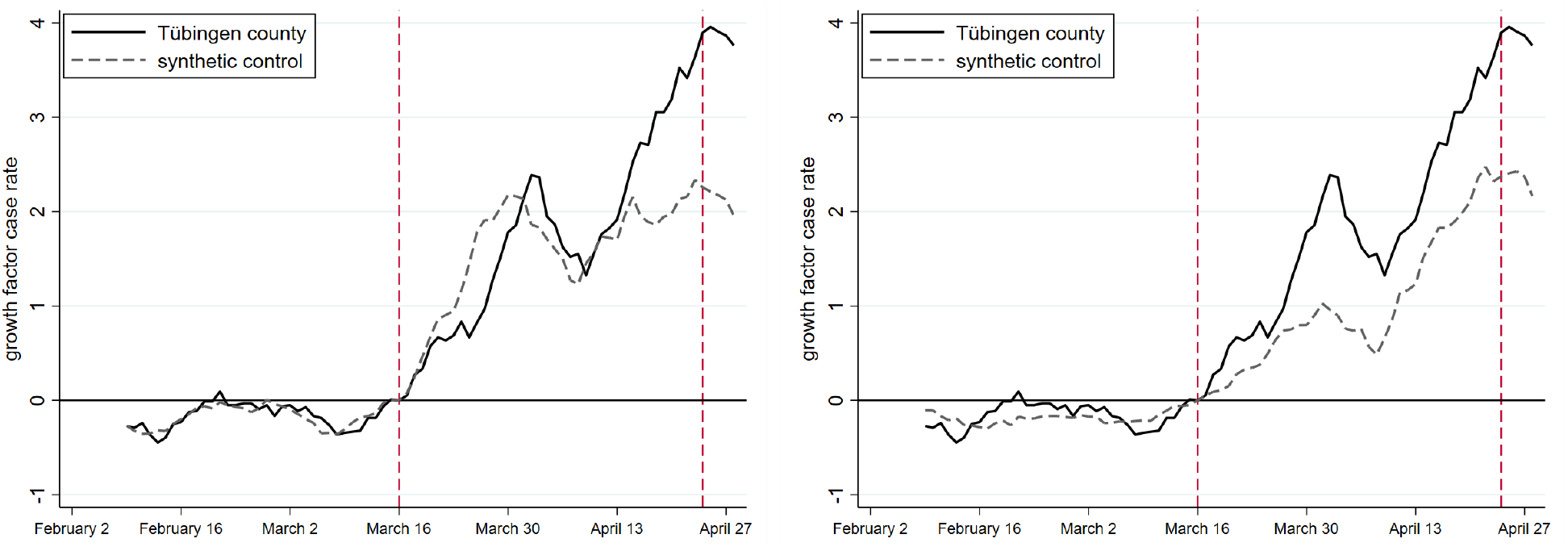
Growth factors for Tübingen county and synthetic control group. Donor pool Germany (left) and Baden-Württemberg (right)

##### Tübingen City

Having shown that replacing the baseline predictor set by a predictor set community and that working with normalized case rates instead of case rates yields the same findings for Tübingen county, we can now study Tübingen city by employing the predictor set community and normalized case rates. The results are in figure 20, the predictor balance table follows thereafter, as does the table with control counties.

**Figure 20:**
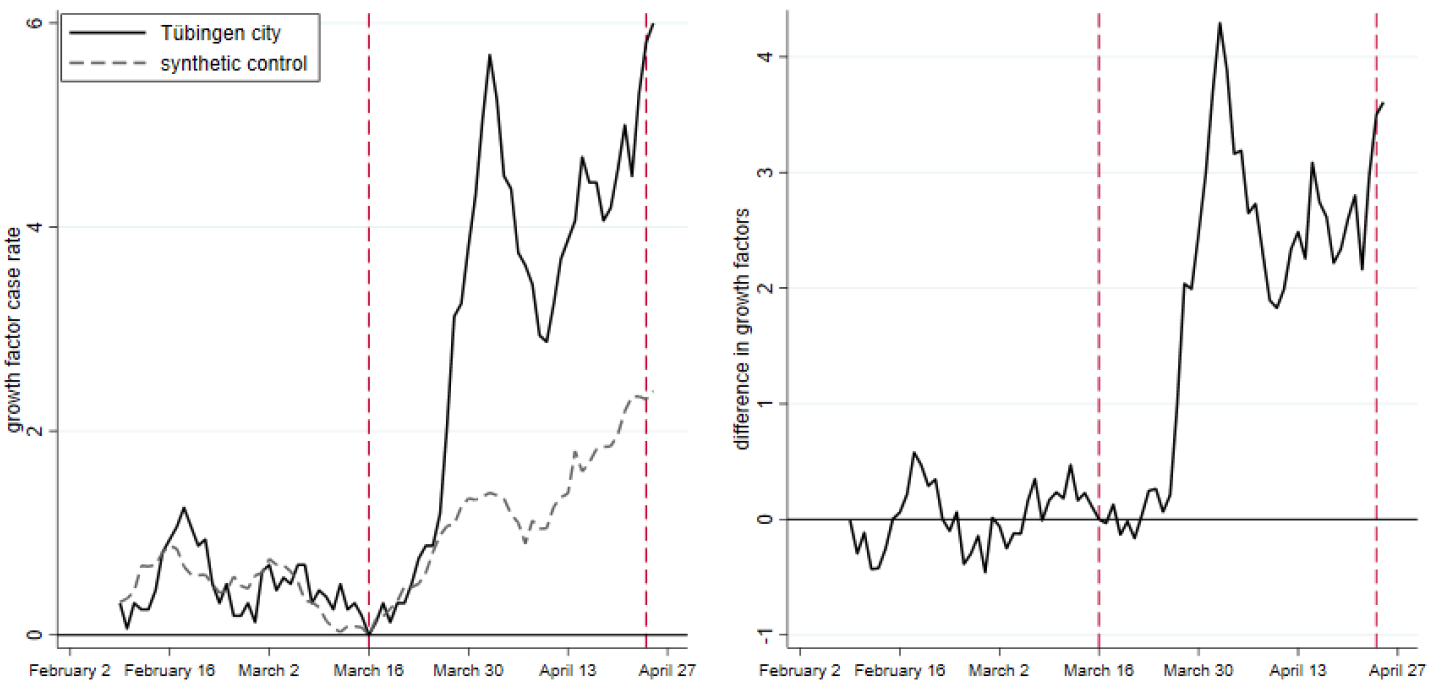
Growth factors for Tübingen city and synthetic control group

This figure clearly shows that the growth in the case rate in Tübingen city is much larger than in the control county. The right panel shows that the increase is higher by a factor of around 3.

We conclude from this that there are effects in Tübingen city as well. The conclusion that the rise in cases in Tübingen county is solely due to an increase in cases outside of Tübingen city would not be correct.

### A.10 The future of Opening under Safety

In case the future of the pandemic requires OuS (we hope not), what do we learn from the Tübingen experience? How do our findings compare to more successful OuS projects as reported in (*37*)?

First, the link between testing centers and health authorities must be strengthened. Positive rapid testing must be PCR confirmed and reported back to testing centers. Data from testing centers would then be much more informative. Second, infection data at the community level should be made public in a systematic way.

Third, data on individuals that participate in OuS projects should be collected. Our evaluation is based on aggregate county and community data. It would be much more informative if we knew whether individuals taking part in OuS are actually infected around 5-7 days after the event they took part in. Such a ‘terminal test’ joint with the initial rapid test would allow to draw conclusions about OuS much more convincingly. If the participation of individuals in individual events was also registered (for some examples, see (*37*)), one could also draw conclusions about individual components of OuS (is going to the cinema more risky than going shopping?).

Comparing OuS in Augustusburg with OuS in Tübingen, some conjectures can be made which, at this point, require more data to be confirmed. This discussion also relates to the ‘catching up’ hypothesis laid out above. Why could ‘catching up’ ever take place? Only when testing is not perfect. It is clear that any testing procedure produces false negatives. It is also clear that not all infected and infectious individuals can be detected by rapid testing (*38*). Hence, testing centers, their management and their personnel is crucial in keeping the real-world (as opposed to laboratory) number of false negatives low. We have no data whatsoever to claim that testing centers were at the basis of the rise of infections in this OuS project. Other hypotheses should be taken into account as well in case future OuS projects will take place and will be evaluated.

## Notes

### Competing Interest Statement

The authors have declared no competing interest.

### Funding Statement

no external funding

